# Mental health symptoms in Latin America during the first year of COVID-19 pandemic: a meta-analysis of prevalence and potential moderator variables

**DOI:** 10.1101/2023.02.18.23286126

**Authors:** F. Torrente, D. Ailán, E. Del Cerro, J. Del Negro, B. Gorodetzky, D. Slonimschik, M. Cetkovich-Bakmas, P. López

**Affiliations:** Institute of Cognitive and Translational Neurosciences, (CONICET-INECO Foundation-Favaloro University)

**Keywords:** depression, anxiety, stress, distress, sleep quality, loneliness, lockdown

## Abstract

This meta-analysis examines the impacts of the COVID-19 pandemic on mental health in Latin America during its first year, using data from 71 studies with 231,441 participants. To address knowledge gaps in the existent literature we considered the type of study design, country of origin, effects of lockdown, and several potential moderating factors. We found prevalence rates of 31% for depression symptoms and 36% for anxiety symptoms. These estimates were higher than those observed in studies from the northern hemisphere. Longitudinal studies showed that depressive symptoms persisted over time, and lockdowns were associated with mildly increased anxiety levels. Female gender was associated with higher rates of depressive symptoms. Our analyses revealed high statistical heterogeneity, and moderating factors such as pandemic duration, COVID-19 cases and deaths, and lockdown stringency did not explain observed mental health symptoms. Methodological limitations include an overreliance on cross-sectional studies and a lack of pre-pandemic parameters that may lead to an overestimation of mental health symptom rates. Overall, this study provides valuable insights into mental health symptoms in Latin America during the first year of the COVID-19 pandemic, highlighting the need for improved epidemiological research and mental health support in the region.

## 1. Introduction

As time passes since the beginning of the COVID-19 pandemic, we are gaining more insight into its various impacts on the mental health of the population worldwide. Several meta-analyses indicated that mental health symptoms increased during the pandemic globally (de Sousa et al., 2021; Phiri et al., 2021; Wu et al., 2021). A meta-analysis of 48 studies analyzing the global prevalence of depressive and anxiety disorders during the COVID-19 pandemic found elevated rates of the two conditions during 2020 compared to a pre-pandemic baseline (Santomauro et al., 2021). Both depression and anxiety were associated with increased SARS-CoV-2 infection rates and decreased human mobility. In a meta-review of meta-analyses, de Sousa et al. (2021) found a prevalence of 28.33% for anxiety symptoms and 26.7% for depression symptoms in the general population.

However, most of these estimates came from studies from the northern hemisphere, and more specifically from the US, Europe, and China. Among the regions with less information on the matter is Latin America, a region where a high impact on mental health was expected, given the socioeconomic inequities, the insufficient health services, and the high rates of infections and mortality due to COVID-19 in some of the countries (Kola et al., 2021). In support of these assumptions, a previous meta-analysis of mental health symptoms during the COVID-19 pandemic in Latin America (Zhang et al., 2022) covering 62 studies showed a pooled prevalence of anxiety, depression, distress, and insomnia of 35%, 35%, 32%, and 35%, respectively, which would indicate higher rates than those reported globally.

Beyond these general estimates, several questions remained unanswered in the Latin American context. Some studies have indicated that prevalence rates in cross-sectional studies are exaggerated compared to longitudinal studies and studies that include pre-pandemic measures. For example, a meta-analysis of studies comparing measurements before and after the onset of the pandemic showed mild increases in mental health symptoms that attenuated over time (Robinson et al., 2022). In contrast, other studies showed more pronounced differences between pre and post-pandemic measures (Santomauro et al., 2021). Also, it is possible that mental health indicators suffered variations over time due to changes in contextual factors (Cénat et al., 2022) and to the action of adaptative mechanisms related to resilience (Fancourt et al., 2021; Killgore et al., 2020). For this reason, it may be important to compare the results of cross-sectional versus longitudinal studies to obtain a more accurate estimate of the impact of the pandemic on mental health in Latin America, since longitudinal studies could more accurately reflect the adaptive response of individuals to stressors and the action of resilience mechanisms over time. The previous meta-analysis of mental health in Latin America (Zhang et al., 2022) does not compare estimates from different types of study, nor does it take into account studies with measurements before the pandemic.

Furthermore, considering the differences within the region in terms of the health and socioeconomic impact of the pandemic, it may be of interest to explore the differences between countries in the rates of mental health symptoms, and also contextual differences in the countries while the studies were carried, for example, whether there was a lockdown during the study period or not. Lockdowns have been shown to influence mental health estimates in previous studies (Prati and Mancini, 2021) and hence it would be of interest to know how they impacted in the context of Latin America.

Finally, the existent meta-analysis of mental health in Latin America during the pandemic (Zhang et al., 2021) did not inform about potential moderators of the prevalence of the symptoms, such as infection and mortality rates, the stringency of lockdowns, or time elapsed since the beginning of the pandemic, as it has been reported in other worldwide meta-analyses (Cénat et al., 2022; de Sousa et al., 2021; Santomauro et al., 2021).

Accordingly, the general aim of this study was to estimate the prevalence of mental health symptoms among the general population of Latin American countries during the first year of the COVID-19 pandemic and to disentangle potential moderators and confounders. For this purpose, we performed a systematic review and meta-analysis of published studies reporting different mental health indicators in the region since the emergence of the COVID-19 pandemic. As specific aims, we first attempted to estimate the pooled global prevalence of mental health symptoms, to be able to compare them with previous meta-analyses in the region and globally. Second, we aimed to compare the results of cross-sectional studies with those of longitudinal studies and to identify the availability of studies that compared measurements before the pandemic with measurements after its onset. Third, we tried to find differences in the prevalence of psychological symptoms according to the country of origin, whether there was a lockdown during the study period or not, and differences between gender groups. We expected that studies performed during the lockdown period would have higher prevalence rates of depression and anxiety symptoms. And fourth, we performed a regression analysis to evaluate potential moderating factors related to mental health symptoms in the region of Latin America. The factors evaluated were the days since the start of the pandemic, COVID-19 cases per million population, deaths due to COVID-19 per million population, and a lockdown index according to the data collection dates of the different studies. In all these cases, we expected that higher values in the moderator variables would be associated with increased prevalence rates of mental health symptoms.

## 2. Methods

We conducted a systematic review and meta-analysis of observational studies reporting the prevalence of mental health symptoms among the general population of Latin American countries during the COVID-19 pandemic. The review protocol was registered on the international prospective register of systematic review (PROSPERO) with registration number CRD42021276277. The Preferred Reporting Items for Systematic Reviews and Meta-Analyses (PRISMA) checklist was followed.

### 2.1. Search strategy and study selection

A comprehensive search of five electronic databases (PubMed, Embase, CINAHL, PsycINFO, LILACS) and two pre-print databases (PsyArXiv, MedRxiv) was conducted to identify observational studies in multiple languages (Spanish, Portuguese, and English), which reported the prevalence of mental health symptoms in the context of COVID-19 since the beginning on the pandemic. We searched all databases from their inception date to September 17, 2021. There was no restriction applied such as ethnicity or publication date.

All articles identified by database searches were uploaded to the systematic review software Covidence (2019). After de-duplication was performed in Covidence, blinded screening at the title-abstract level was completed by four reviewers for inclusion in the full-text review. Further screening of full texts was done by two reviewers to include or exclude based on pre-established criteria. Any disagreement during the selection process was resolved either through consensus or through discussion with a third reviewer.

### 2.2. Eligibility Criteria

We included studies if they (1) were observational studies; (2) reported the prevalence of psychological symptomatology among the general population (18 years+) of Latin America during the COVID-19 pandemic; (3) used standardized and validated measurement instruments to assess mental health outcomes. Full-text articles and preprints were included. We excluded studies if they (1) were interventional studies; (2) were systematic reviews; (3) focused exclusively on healthcare professionals, patients with a current COVID-19 diagnosis, or children; (4) had unclear methods.

### 2.3. Outcomes

The main outcomes considered were anxiety symptoms, depression symptoms, sleep problems, and stress. Additional outcomes were psychological well-being, substance abuse, loneliness, and disturbances of circadian rhythms and/or sleep.

### 2.4. Data extraction

Two reviewers extracted data from all the final studies using a standardized data extraction form, which adheres to the Preferred Reporting Items for Systematic Reviews and Meta-Analysis Protocols (PRISMA-P) 2015 statement (Moher et al., 2015). The following data were extracted:

1. General information: study title, author, publication year, country.
2. In the Methods section: study design, study setting, study participants, sample size, diagnostic tool, and statistical tests employed.
3. In the Outcome section: mean age, non-response rate, the prevalence of stress, anxiety symptoms, depressed mood, sleep quality, and insomnia.

### 2.5. Risk of bias (quality) assessment

The risk of bias was assessed by applying Strengthening The Reporting of Observational Studies in Epidemiology (STROBE), which was designed to ensure a clear presentation of what was planned, done, and found based on the design, participants, sample size, measurement tools, data collection methods, confounders, and statistical analysis. Two reviewers independently rated the 22 items of the STROBE checklist for each article and achieved consensus through discussion. Discrepancies were settled by a third author. Items were scored as 1= reported, 0= not reported.

### 2.6. Statistical analysis

Meta-analysis was performed using Stats Direct. For each of the studies, the standard error was calculated using the reported number of outcomes and the sample size. The Freeman-Tukey double arcsine transformation was used to stabilize the variance of the raw data. We performed a random-effects model to obtain pooled prevalence estimates, which give greater weight to studies with larger sample sizes, with associated 95% confidence intervals (CIs). The *I²* statistic was used to assess the statistical heterogeneity between studies. Statistical heterogeneity was considered present when *p* < 0.1 or *I²* > 50%. Egger regression and funnel plots were used to assess publication bias. Statistical significance was set at *p* < .05. Sensitivity analyses were conducted to evaluate the impact of the methodological quality of included studies on pooled results and to explore potential sources of heterogeneity.

We performed subset analysis for different groups by sex, country, lockdown, and type of study. Cross-country comparisons were limited to four countries due to the scarce number of studies in most countries. Additionally, in the protocol, we had planned to perform subgroup analysis for age groups and waves of COVID-19. However, we have not been able to carry out these analyses due to differences in the intervals of the age groups in the different papers and because we could not find a clear official consensus regarding the start and end dates of each wave of COVID-19 in the different countries of Latin America.

Finally, we carried out a regression analysis with the prevalence of depressive, anxiety, and stress symptoms as dependent variables and cases of COVID-19 per million inhabitants, deaths per million inhabitants, and lockdown stringency index as independent variables. The three indicators for the countries of origin for each of the selected studies were obtained from the COVID-19 Data Explorer of Our World in Data (Ritchie et al., 2020).

## 3. Results

### 3.1. Studies included

Our database searches returned 2893 records (2884 unique records). We added 2 papers by handmade search. After screening titles and abstracts, we deemed 2576 records to be irrelevant and retrieved 310 full texts for further scrutiny. Of these 310 reports, 71 studies met our predefined inclusion criteria. PRISMA diagram is presented in Figure 1.

**Figure 1.**
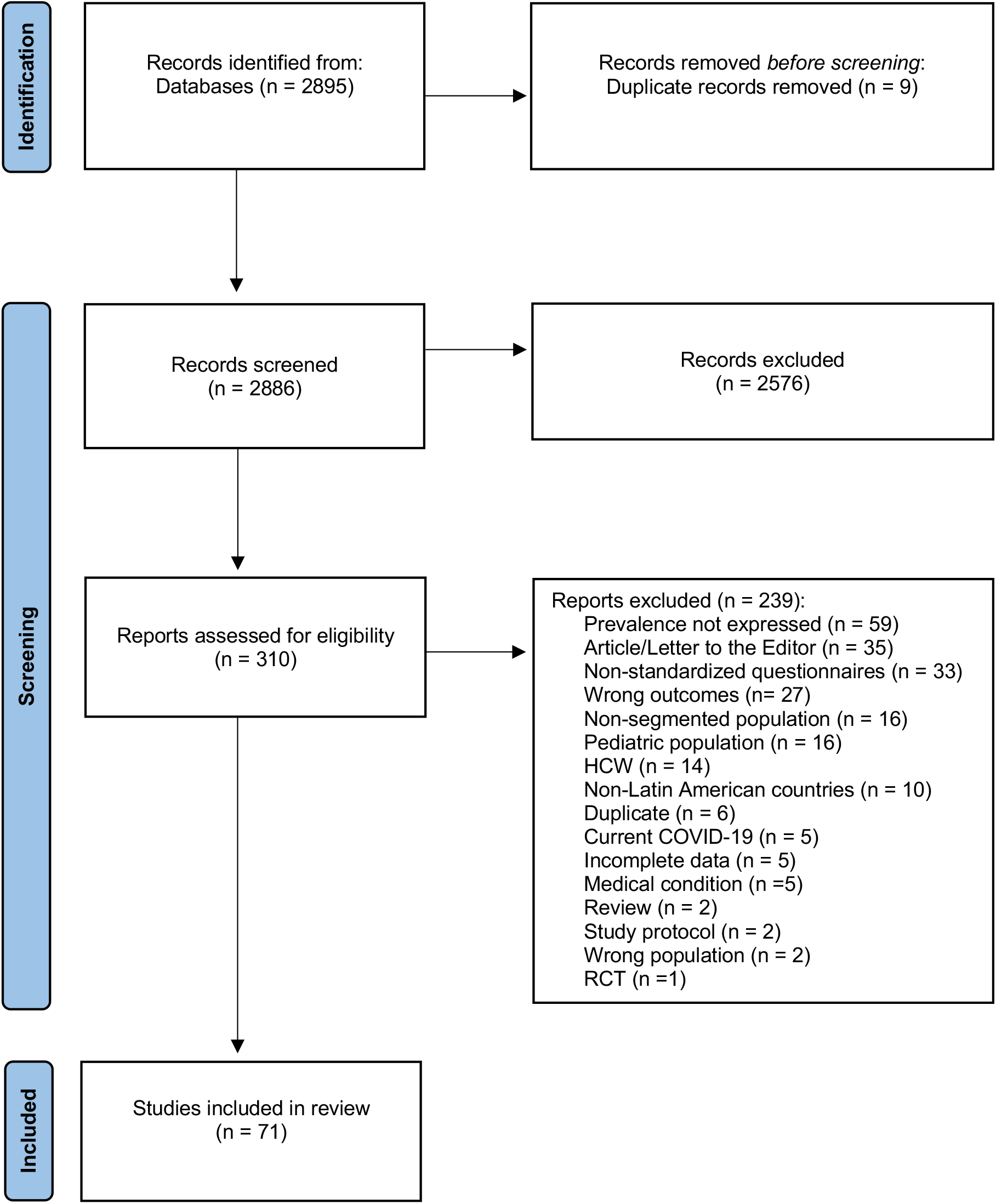
PRISMA Diagram.

### 3.2. Characteristics of included studies

Included studies covered a total of 231441 participants, with sample sizes ranging from 57 to 57250. No population-representative studies were retrieved. All studies reported data from convenience samples. Selected studies came from Argentina, Brazil, Chile, Colombia, Cuba, Ecuador, México, Paraguay, and Perú. An additional study surveyed 15 countries from Latin America and the Caribbean. All included surveys were conducted between March and December 2020.

Ten studies were longitudinal, collecting data at least at two time points. Of them, seven studies collected all the data during the pandemic, while three studies compared measures taken during the pandemic with a pre-pandemic measurement.

Instruments employed for measuring mental health symptoms were highly diverse. The most common instrument used for assessing depression symptoms was the PHQ-9 (16 studies) and the GAD-7 for anxiety symptoms (15 studies).

Details of the 71 studies are presented in Table S1 in Supplementary Material.

### 3.3. Risk of bias of included studies

In the assessment of the risk of bias, the studies addressed 79% of the STROBE checklist items, on average. The four items that were addressed by 100% of the articles were reporting of background/rationale, information of variables, data sources/measurement, and the interpretation of results. However, no articles met all guidelines for the reporting of information. The common issues were failure to report (1) descriptive data (85.9%, n= 61), (2) statistical methods (80.2%, n= 57), and (3) any efforts to address potential sources of bias (76%, n= 54) (See details in Table 1).

**Table 1.**
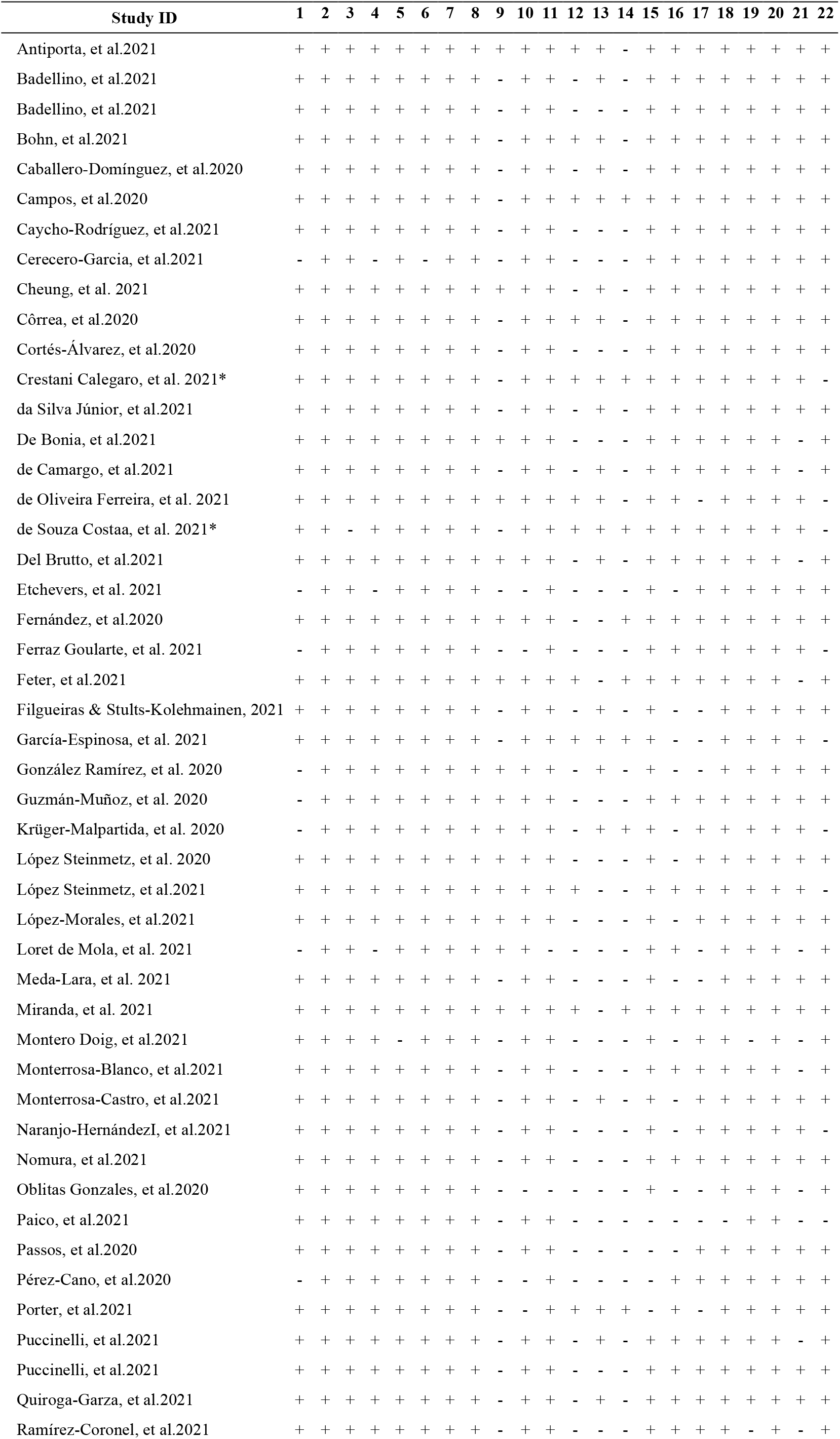
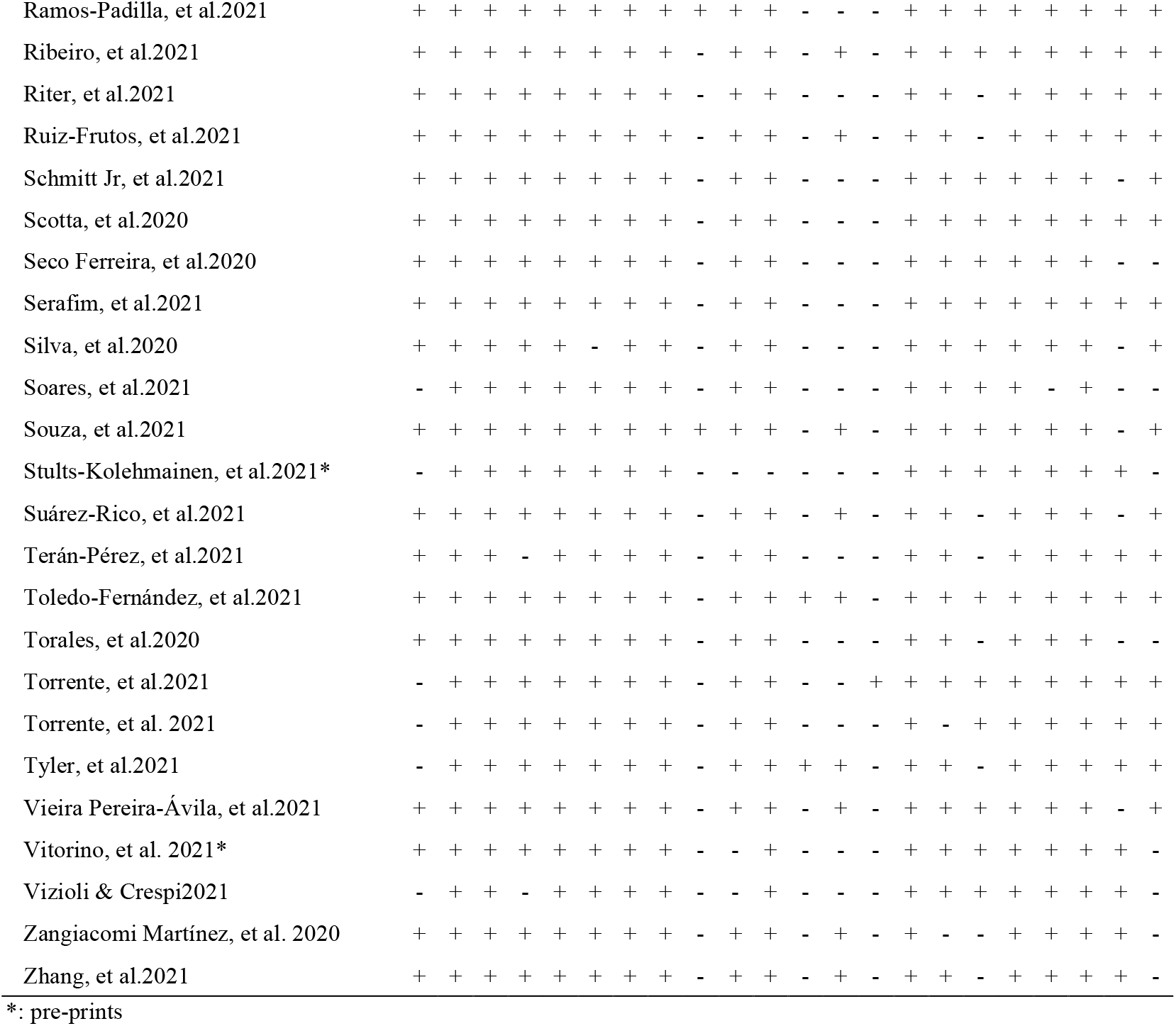
Risk of bias in included studies – Strobe Checklist.

### 3.4. Prevalence of depressive symptoms

The pooled prevalence of depressive symptoms was 30.99% (95% CI = 26.83-35.32%; see Figure 2) in the 47 studies included in this analysis. Heterogeneity was found to be high (*I²* = 99.6%). In all the studies that reported depressive symptoms segregated by sex, a higher prevalence was observed in women than in men (See Table 2).

**Table 2.** Prevalence of moderate to severe depressive, anxiety, and stress symptoms according to gender.

|  | Female | Male |
| --- | --- | --- |
| *Depressive symptoms* |  |  |
| Antiporta et al. 2021 | 35.3% | 23.7% |
| Badellino, et al. 2021 | 51.6% | 36% |
| Puccinelli, et al. 2021 | 35.3% | 21% |
| Silva, et al. 2020 | 60.4% | 40.8% |
| Terán-Pérez, et al. 2021 | 24.5% | 18.6% |
| Vieira Pereira-Ávila, et al. 2021 | 10.3% | 5.9% |
| *Anxiety symptoms* |  |  |
| Filgueiras & Stults-Kolehmainen 2021 | 8.9% | 8.4% |
| Oblitas Gonzales, et al. 2020 | 3.7% | 27.5% |
| Puccinelli, et al. 2021 | 28.3% | 15.5% |
| Silva, et al. 2020 | 46.4% | 28.7% |
| Terán-Pérez, et al. 2021 | 36.3% | 35.9% |
| *Stress symptoms* |  |  |
| de Camargo, et al. 2021 | 21.3% | 42.8% |
| González Ramírez, et al. 2020 | 30% | 20% |
| Silva, et al. 2020 | 51.74% | 31.61% |

**Figure 2.**
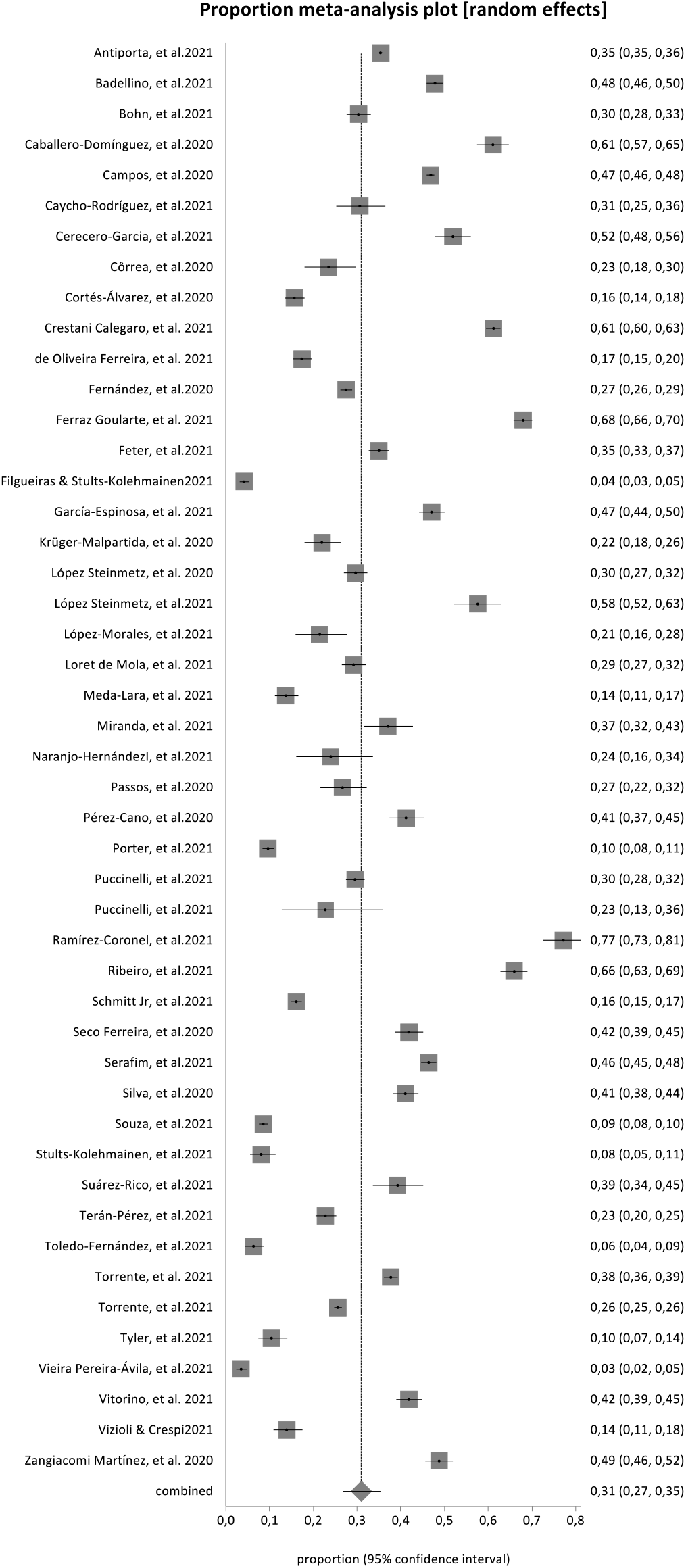
Pooled prevalence of depression symptoms in all studies included.

Regarding the type of study design, the pooled prevalence of depressive symptoms in longitudinal studies was 29.68% (95% CI = 15.93 to 45.64%, *I*^*2*^ = 99.3%; see Figure 3), while the prevalence observed in cross-sectional studies was 31.22% (95% CI = 26.73 to 35.89%, *I*^*2*^ = 99.6%; see Figure S1 in Supplementary material). One study reported a 5.7-fold increase in depression symptoms rate during the COVID-19 pandemic compared to pre-pandemic measurements (from 5.1 to 29.3%; see Table S1, Loret de Mola et al., 2021).

**Figure 3.**
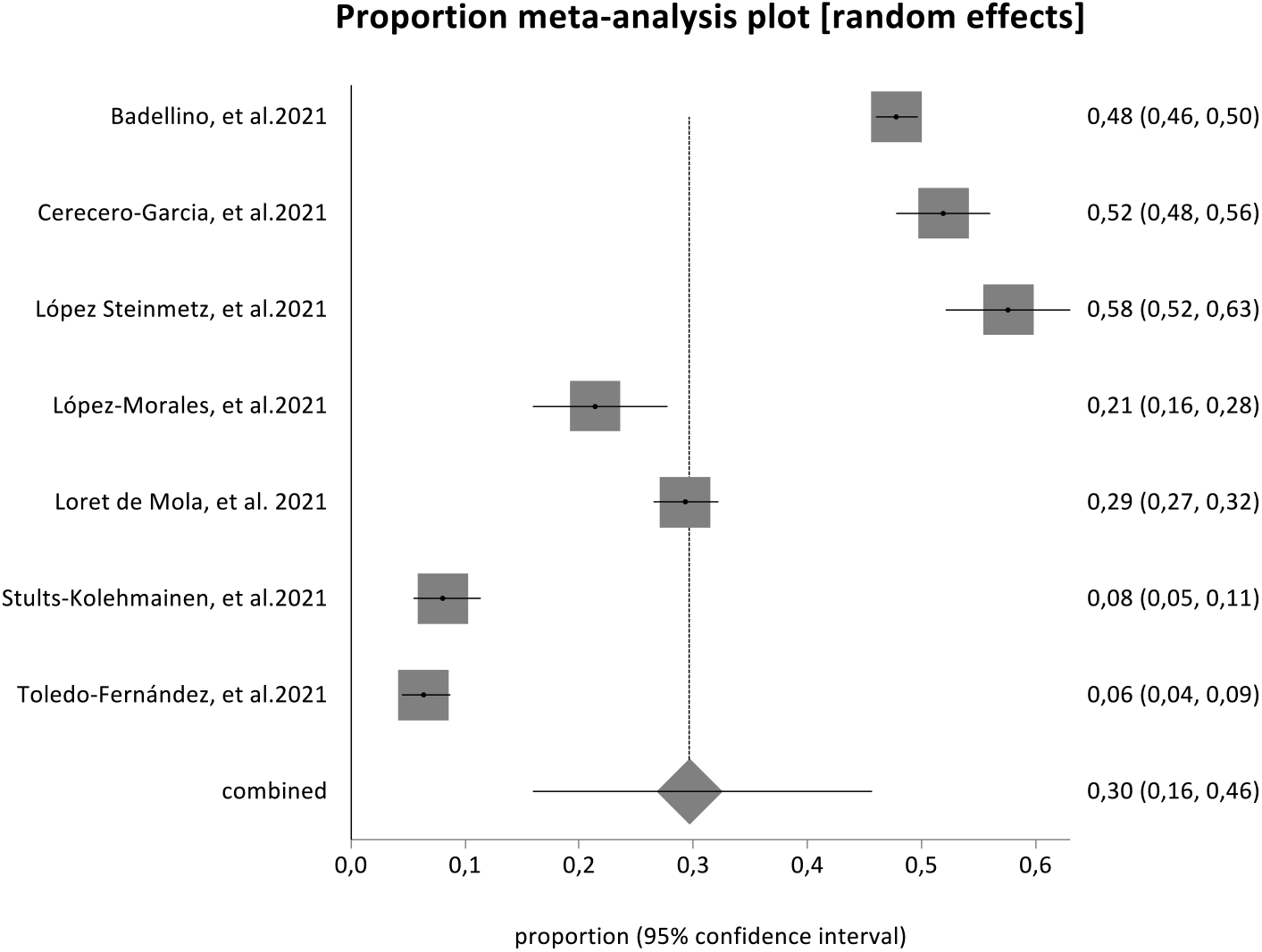
Pooled prevalence of depression symptoms in longitudinal studies.

The pooled prevalence of depressive symptoms in studies carried out during lockdowns (pooled prevalence = 28.63%, 95% CI = 24.17 to 33.31%; *I*^*2*^ = 99.5) was similar to that observed in studies without lockdown (pooled prevalence = 27.37%, 95% CI = 23.75 to 31.14%; *I*^*2*^ = 75.8%).

Sub-analysis by country of origin showed a higher prevalence of depressive symptoms in Argentina (9 studies; pooled prevalence = 32.61%, 95% CI = 26.23 to 39.33%; *I*^*2*^ = 99%) and Brazil (22 studies; pooled prevalence = 30.83%, 95% CI = 22.60 to 39.72%; *I*^*2*^ = 99.7%) in comparison with Mexico (7 studies; pooled prevalence = 25.65%, 95% CI = 14.85 to 38.23%; *I*^*2*^ = 98.9%) and Peru (4 studies; pooled prevalence = 23.59%, 95% CI = 9.34 to 41.86%; *I*^*2*^ = 99.6%) (See Figure S2 in Supplementary material).

### 3.5. Prevalence of anxiety symptoms

The pooled prevalence of anxiety symptoms in the 43 studies included in this analysis was 35.94% (95% CI = 30.51-41.55%, see Figure 4). Heterogeneity was high (*I*^*2*^ = 99.6%).

**Figure 4.**
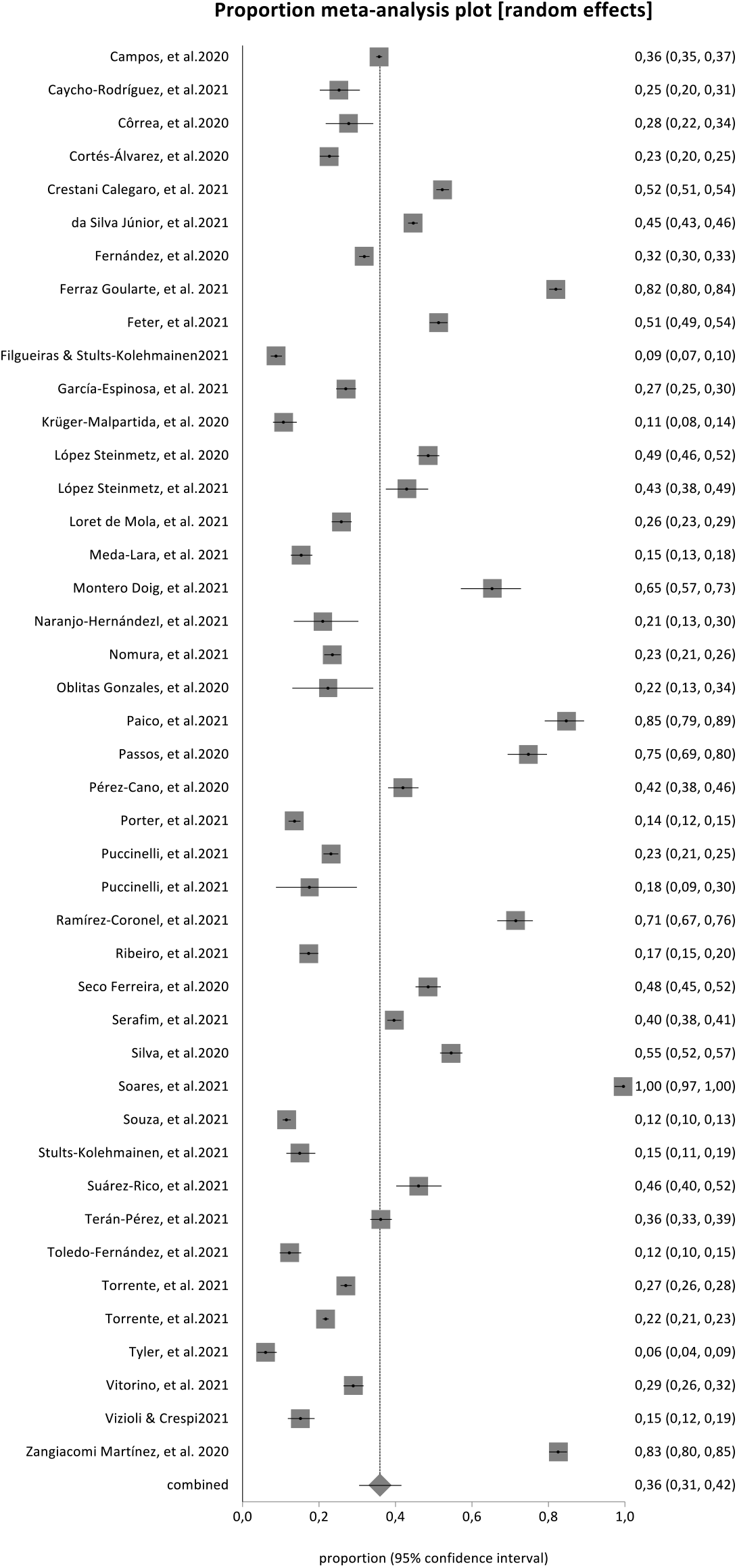
Pooled Prevalence of anxiety symptoms in all studies included.

**Figure 5.**
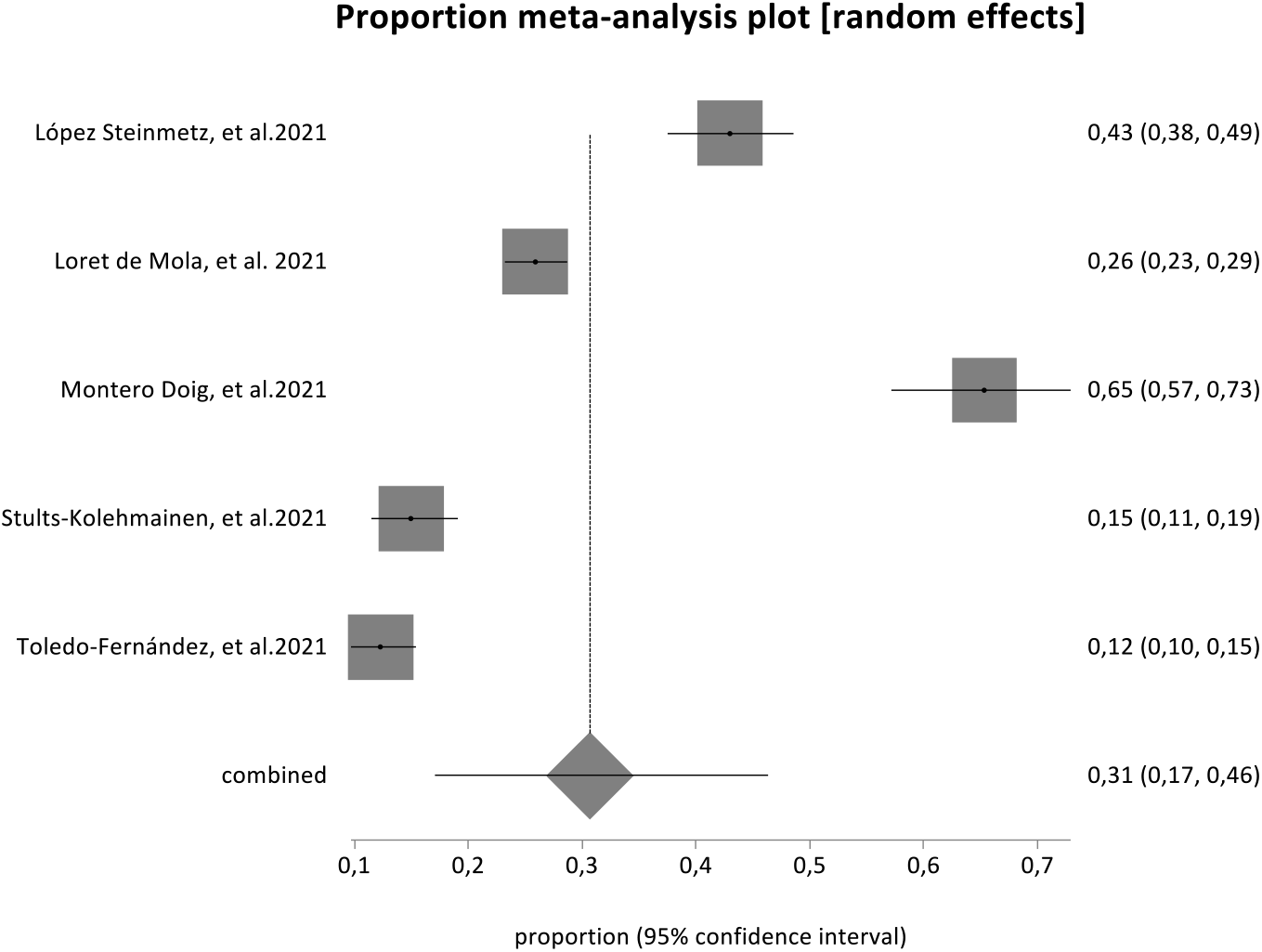
Pooled prevalence of anxiety symptoms in longitudinal studies.

Five studies presented results disaggregated by sex. Two studies showed higher prevalence rates of anxiety symptoms in women than in men, one study presented higher prevalence in men, and two studies showed similar rates (See Table 2).

The prevalence of anxiety symptoms was lower in longitudinal studies (pooled prevalence = 30.69%, 95% CI = 17.08 to 46.28%, *I*^*2*^ = 98.3%; see Figure 4) in comparison with cross-sectional studies (pooled prevalence = 36.63%, 95% CI = 30.82 to 42.65%; *I*^*2*^ = 99.6%; see Figure S3). One small study in Perú (n=150) with a pre-pandemic baseline showed increased anxiety during the pandemic (a 26.6% increase in cases of moderate and severe anxiety; see Table S1, Montero Doig et al., 2021).

Studies carried out during lockdown (pooled prevalence = 33.52%, 95% CI = 27.37 to 39.96%; *I*^*2*^ = 99.4%) showed a higher prevalence of anxiety symptoms than studies implemented in periods without lockdown (pooled prevalence = 17.06%, 95% CI = 8.55 to 27.74%; *I*^*2*^ = 94.6%).

Sub-analysis by country of origin (see Figure S4 in Supplementary Material) showed higher anxiety symptoms prevalence in Brazil (21 studies; pooled prevalence = 41.26%, 95% CI = 32.54 to 50.26%; *I*^*2*^ = 99.7%) and Peru (6 studies; pooled prevalence = 35.68%, 95% CI = 14.84 to 59.87%; *I*^*2*^ = 99.2%), compared to Argentina (6 studies; pooled prevalence = 30.56%, 95% CI = 23.61% to 37.98%, *I*^*2*^ = 99%) and Mexico (6 studies; pooled prevalence = 28.08%, 95% CI = 18.35 to 39.00%; *I*^*2*^ = 98.3%).

### 3.6. Prevalence of other mental health symptoms

The pooled prevalence of stress symptoms corresponding to the 18 studies included in this analysis was 41.20% (95% CI = 31.00 to 51.79%; *I*^*2*^ = 99.7%; see Figure S5). In one study men presented a higher prevalence of stress symptoms than women (See Table 2)

The pooled prevalence was 38,62% in 7 studies of distress (95% CI = 29.86 to 47.77%; *I*^*2*^= 99.2%; see Figure S6), 53.49% in 6 studies of sleep quality (95% CI = 41.35 to 65.43%, *I*^*2*^ = 99.4%) and 38.19% in 3 studies of loneliness (95% CI = 26.03 to 51.15%, *I*^*2*^ = 98.4%; see Figure S7).

For the other variables reported in the included studies, such as sleep-related measures other than sleep quality, suicide, alcohol consumption, and quality of life, the pooled prevalence was not calculated because there were not enough studies to calculate pooled prevalence.

Finally, one study with pre-pandemic baseline estimates showed increased sleep problems (49% of participants were categorized as “poor sleepers” in comparison with 29% of participants at baseline; see Table S1, Del Brutto, et al.2021).

### 3.7. Regression analysis

We explored potential moderator factors for depressive symptoms, anxiety symptoms, and stress through multiple regression models. The explanatory factors evaluated were the days since the start of the pandemic, COVID-19 cases per million population, deaths due to COVID-19 per million population, and a lockdown index according to the data collection dates of the different studies. None of the variables included in the regression analyses were significant predictors for any of the three dependent variables (see Supplementary Material for further details).

## 4. Discussion

In this meta-analysis of 71 studies, we aimed to revise and synthesize the evidence about mental health symptoms during the COVID-19 pandemic in Latin America considering the type of design, country of origin, lockdown effects, and several potential moderating factors. The first aim was to know the general pooled prevalence of the different mental health symptoms. We found a pooled prevalence of 31% for depression and 36% for anxiety symptoms. Besides anxiety and depression, we observed a pooled prevalence of 41% for stress symptoms, 39% for distress, 53 for problems in sleep quality, and 38% for loneliness. In comparison with the former meta-analysis of mental health in Latin America during COVID-19 (Zhang et al., 2022) our estimates were slightly lower for depressive symptoms (31% vs. 35%) and similar for anxiety symptoms (36% vs. 35%). Both regional revisions obtained higher rates than those reported by de Sousa et al. (2021) in their global meta-review (27% for depression and 28% for anxiety symptoms). The differences in rates with other regions may be attributed to socioeconomic, health, and contextual factors, or even to political factors, such as trust in the government and the healthcare system (Kola et al., 2021). However, we cannot discard methodological reasons for these elevated prevalence rates, as we will discuss below.

As a second aim, we sought to compare the results of cross-sectional studies with those of longitudinal studies and looked for studies that compared measurements before the pandemic with measurements after its onset. We found seven longitudinal studies that reported two or more time point assessments during the pandemic and three that compared measures taken during the pandemic with a pre-pandemic baseline. We found that rates of anxiety symptoms were lower in extended measurements of longitudinal studies in comparison with rates observed in cross-sectional studies. This difference may indicate an adaptive process of habituation to the threat, or an effect of the measures taken to combat the spread of COVID-19 that may have increased the population’s perceptions of safety. In contrast, we found no difference between the prevalence rates of longitudinal versus cross-sectional studies for depression, which remained equally high. These results could indicate that the effects of the pandemic on mood symptoms in Latin America may have extended over time, at least during the first year, unlike what was observed in other regions (Robinson et al., 2020). The prolonged impact of the pandemic on mood in the Latin American region could be associated with a sustained perception of disruption of normality, losses of different kinds, economic burden, or mental fatigue (Torrente et al., 2022). However, it should be noted that the low number of longitudinal studies retrieved, the small sample sizes, and the wide confidence intervals observed limit confidence in these interpretations.

In addition, the possibility of estimating actual changes in mental health symptomatology after the start of the pandemic becomes uncertain because of the lack of comparative pre-pandemic baseline data obtained with reliable standardized measures from the same population. As we observed in the present meta-analysis, only three studies were retrieved in our search that compared estimations before and after the beginning of the pandemic, showing increases in mental health symptoms (depression, anxiety, and sleep problems). However, two of the studies were small sample studies, two were focused on specific populations, and one evaluated only sleep problems. The lack of more and larger studies comparing estimates during the pandemic with pre-pandemic parameters appears as a severe deficit in the published literature on mental health in Latin America during the COVID-19 pandemic.

Our third aim was to evaluate the influence of different factors in the prevalence rates of mental health symptoms, such as country of origin, presence of a lockdown, and gender. Despite the small number of nations scoped in the retrieved studies, the regional sub-analysis revealed some disparities, with Brazil and Argentina having greater rates of depressive symptoms, and Brazil and Peru having higher rates of anxiety. Also, we found that studies conducted during periods of lockdown registered higher levels of anxiety than those that were not, although we did not find an association through regression analyses between the level of mental health estimates and the index of lockdown stringency. Confinement periods are associated with the higher circulation of the virus, more infections, and before vaccines, with more mortality, so it is plausible that this context of increased threat translated into higher levels of anxiety. This result is also consistent with the mild increase in anxiety reported in a previous meta-analysis about the impact of lockdowns during COVID-19 (Prati and Mancini, 2021). However, the observed association in our review should be interpreted with caution because the comparison was based on only four studies without lockdown. In contrast, we observed no association between increased symptoms of depression and confinement. This outcome was against our expectations since in previous literature it was found that lockdowns were also associated with mild increases in depression (Prati and Mancini, 2021). Among the contextual factors that might have played a role in reducing depression symptoms during lockdowns are citizens’ positive attitudes toward lockdown measures, social support, and financial assistance. Also, individuals’ protective behaviors may have attenuated the impact of lockdowns. Once again, the low number of studies without confinement may have made our comparison insufficiently powered or representative.

Additionally, we found that the prevalence of depressive symptoms was higher in women than in men in all the studies that reported data segregated by gender. Although it is a well-known fact that women have an increased prevalence of depression and anxiety disorders even before the pandemic (Rehm and Shield, 2019), contextual factors could have contributed to their higher prevalence in the present circumstances (Almeida et al., 2020; Thibaut and van Wijngaarden-Cremers, 2020). Daily stress may have been incremented for many women in charge of household chores and childcare in parallel with home-based work. Increased domestic violence may have also contributed to the burden of mental health for women during the confinement periods (Piquero et al., 2021)

Finally, we intended to evaluate potential moderating factors related to mental health symptoms in the region. For this purpose, we carried out a regression analysis with the prevalence of depressive, anxiety, and stress symptoms as dependent variables and cases of COVID-19 per million inhabitants, deaths per million inhabitants, and lockdown stringency index as independent variables. Nevertheless, none of the variables introduced in the analyses showed to be of help for understanding the variance of outcomes in mental health symptoms in the present review. Alongside methodological disparities between the revised studies that may have contributed to the unexplained heterogeneity of results and that we will discuss in the Limitations section, a genuine variation in mental health expressions may have resulted from factors not accounted for in the present study, such as social and cultural attitudes, ethnic differences, healthcare resources, economic context, and government response, among others. As an alternative, significant heterogeneity may reflect the substantial inter-individual diversity in people’s responses to acute stress.

### 4.1. Limitations

The present meta-analysis has several limitations. First, all the performed analyses revealed a high statistical heterogeneity (I² greater than 75%) that was not explained by recognizable variables. Several methodological factors may have contributed to this high unaccounted heterogeneity, including the use of diverse instruments to measure mental health symptoms, variations in cut-offs or criteria for classifying relevant conditions, and reliance on convenience sampling (Pierce et al., 2020). Second, as we discussed before, the preponderance of cross-sectional studies in our review may have overestimated the real prevalence rates of mental health symptoms in the region, as was also the case in the previous regional meta-analysis (Zhang et al., 2021). Third, all the reported rates were based on self-reported instruments which may also overestimate the rate of psychiatric disorders in comparison with the more reliable gold standard of diagnostic interviews. Fourth, the period covered by the included studies corresponds to 2020, which restricts the possibility of observing longer adaptation phenomena and does not allow the long-term impact of the pandemic on mental health to be assessed. Fifth, despite a comprehensive search strategy, data on different countries were limited, with many countries of the region missing and others overrepresented, and the reporting of outcomes other than depression and anxiety was limited to a small number of studies, and in some important cases, such as suicidal ideation or substance abuse, there were not enough reports to calculate the pooled prevalence. Sixth, it was not possible to do a subgroup analysis based on the age group and waves of Covid-19 as planned, because they were no consistent data or criteria. In addition, other potentially relevant characteristics, such as economic indicators or public health resources, were not included in the analysis.

### 4.2. Implications and further directions

As we discussed previously, the literature on the prevalence of mental health symptoms in Latin America suffers from important gaps. The most important of the shortcomings is the lack of prior parameters to estimate the true impact of the pandemic against a reliable baseline. This gap is due to the scant epidemiological research on mental health in the region. Also, none of the studies included in this meta-analysis reported the results of a population-representative cohort, nor the results of a large prospective study started before the pandemic. Therefore, an important implication of this review is the urgent need to develop research focused on the sustained measurement of mental health indicators in the region through representative cohort designs.

Despite these limitations, the results of this review showed an elevated impact of the pandemic on mental health in the short term that could have extended over time more in Latin America than in other regions. This could be due to socioeconomic, health, or political factors. As an implication of this finding, the preparation for possible new pandemics in this region must consider an organized response to the mental health challenges that these types of events entail. As a matter of further study, it would be of interest to know how mental health indicators evolved in the region after the period covered by this review and in particular to analyze the results of the longitudinal studies that may have extended beyond the first year of the pandemic.

### 4.3. Conclusions

Our meta-analysis based on 71 studies revealed that the prevalence rates of depression and anxiety symptoms in Latin America were higher than global estimates based on studies from the northern hemisphere. Also, longitudinal studies have shown that the elevated rates of depressive symptoms persisted over time, at least during the first year of the pandemic. Lockdowns appear to be associated with mildly increased anxiety levels. Being female was associated with higher rates of depressive symptoms. The observed rate of depression, anxiety, and stress symptoms in the general population could not be explained by moderating factors such as days since the start of the pandemic, COVID-19 cases per million population, deaths due to COVID-19 per million population, and lockdown stringency index. The interpretation of these results must be cautious due to the methodological limitations of the reviewed literature as a whole. To remedy these deficiencies and to prepare for future pandemics or similar stressful events, there is an urgent need to develop solid epidemiological research on the evolution of indicators of mental health in Latin America.

## Data Availability

All data produced in the present work are contained in the manuscript and in the Supplementary material

## Acknowledgments

We thank Dr. Agustín Ciapponi and Mg. Daniel Comandé from the IECS (Institute for Clinical Effectiveness and Health Policy, Buenos Aires, Argentina) for their thoughtful guidance during the review process.

## Funding sources

This study was supported by the Interamerican Development Bank (RG-T3106) and INECO Foundation.

## Conflicts of interest

None of the authors declare any competing interests.

## Notes

### Competing Interest Statement

The authors have declared no competing interest.

