## Supplementary Material for "Mental health symptoms in Latin America during the first year of COVID-19 pandemic: a meta-analysis of prevalence and potential moderator variables"

**This PDF file includes:**

Search strategy

Table S1 to S4

Figures S1 to S8

References of included studies

**Search strategy**

(SARS-CoV-2[Mesh] OR COVID-19[Mesh] OR Corona Virus[tiab] OR COVID-19[tiab] OR COVID19*[tiab] OR 2019-nCoV[tiab] OR SARS-CoV-2[tiab] OR SARS-CoV2[tiab] OR (Pneumonia[tiab] AND Wuhan[tiab] AND 2019[tiab]) OR Coronavir*[tiab] OR Coronovir*[tiab] OR Virus Corona[tiab] OR Corono Virus[tiab] OR HCov*[tiab] OR CV19*[tiab] OR CV-19[tiab] OR N-Cov[tiab]) AND (Mental Health[Mesh] OR Mental Health[tiab] OR Depressive Disorder[Mesh] OR Depressi*[tiab] OR Sadness[tiab] OR Anxiety[Mesh] OR Anxiousness[tiab] OR Anxiet*[tiab] OR Distress[tiab] OR Angst[tiab] OR Nervousness[tiab] OR Stress Disorders, Traumatic[Mesh] OR Stress*[tiab] OR Sleep Initiation and Maintenance Disorders[Mesh] OR Insomn*[tiab] OR Sleep*[tiab] OR Early Awak*[tiab] OR DIMS[tiab]) AND (Americas[MeSH Terms:noexp] OR America*[tiab] OR Latin America[Mesh] OR Latin America*[tiab] OR Latinamerica*[tiab] OR Latinoamerica*[tiab] OR Latin*[tiab] OR Hispanic Americans[Mesh] OR Hispanic America*[tiab] OR Hispanoamerica*[tiab] OR Iberoamerica*[tiab] OR Ibero Americ*[tiab] OR Panamerican*[tiab] OR Central America[Mesh] OR Central America*[tiab] OR Centroamerica*[tiab] OR Mesoamerica*[tiab] OR Meso America*[tiab] OR Middle America*[tiab] OR South America[Mesh] OR South America*[tiab] OR Southamerica*[tiab] OR Sudamerica*[tiab] OR “America del sur”[tiab] OR Caribbean Region[Mesh] OR Caribbean[tiab] OR Caribe*[tiab] OR West Indies[Mesh] OR West Indi*[tiab] OR Antill*[tiab] OR Indians, South American[Mesh] OR Indians, Central American[Mesh] OR Amerindian*[tiab] OR Indians[tiab] OR American Indian*[tiab] OR Native America*[tiab] OR Patagoni*[tiab] OR Andes[tiab] OR Andean*[tiab] OR Amazon*[tiab] OR Argentin*[ad] OR Argentin*[tiab] OR Argentina[pl] OR Bolivia*[ad] OR Bolivia*[tiab] OR Bolivia[pl]

OR Brazil*[ad] OR Brasil*[ad] OR Brazil*[tiab] OR Brasil*[tiab] OR Brazil[pl] OR Colombia*[ad] OR Colombia*[tiab] OR Colombia[pl] OR Chile*[ad] OR Chile*[tiab] OR Chile[pl] OR Ecuador*[ad] OR Ecuator*[ad] OR Ecuador*[tiab] OR Ecuador[pl] OR Guiana*[ad] OR Guiana*[tiab] OR French Guiana[pl] OR Guyan*[ad] OR Guyan*[tiab] OR Guyana[pl] OR Paraguay*[ad] OR Paraguay*[tiab] OR Paraguay[pl] OR Peru*[ad] OR Peru*[tiab] OR Peru[pl] OR Surinam*[ad] OR Surinam*[tiab] OR Suriname[pl] OR Uruguay*[ad] OR Uruguay*[tiab] OR Uruguay[pl] OR Venez*[ad] OR Venez*[tiab] OR Venezuela[pl] OR Belize*[ad] OR Belize*[tiab] OR Belize[pl] OR Costa Ric*[ad] OR Costarric*[ad] OR Costaric*[ad] OR Costa Ric*[tiab] OR Costarric*[tiab] OR Costaric*[tiab] OR Costa Rica[pl] OR Salvador*[ad] OR Salvador*[tiab] OR El Salvador[pl] OR Guatemal*[ad] OR Guatemal*[tiab] OR Guatemala[pl] OR Hondur*[ad] OR Hondur*[tiab] OR Honduras[pl] OR Nicaragu*[ad] OR Nicaragu*[tiab] OR Nicaragua[pl] OR Panam*[ad] OR Panam*[tiab] OR Panama[pl] OR Mexico[Mesh] OR Mexic*[ad] OR Mexic*[tiab] OR Mejic*[tiab] OR Mexico[pl] OR Cuba*[ad] OR Cuba*[tiab] OR Cuba[pl] OR Dominic*[ad] OR Dominic*[tiab] OR Dominican Republic[pl] OR Haiti*[ad] OR Haiti*[tiab] OR Haiti[pl] OR Jamaic*[ad] OR Jamaic*[tiab] OR Jamaica[pl] OR Puerto Rico[Mesh] OR Puerto Ric*[tiab] OR Puertorric*[tiab] OR Puertoric*[tiab])

#### Table S1. Description of included studies

| | Authors | Country | Data collection period | N | Inclusion criteria | Sampling | Assessment tools | Design | Time points | Results | Lockdown |
|---|---|---|---|---|---|---|---|---|---|---|---|
| 1. | Antiporta, et al.2021 | Peru | 5/4/2020 - 5/18/20 | Total sample: 64493. Final sample: 57250 | 18 years or older | Convenience | PHQ-9 | Cross-sectional | - | Depressive symptoms: Participants with previous mental health diagnosis 59% (95%, CI 56.7, 61.4%) / Adults without a prior diagnosis 30% (95% CI 29.1, 30.9%) / Non-responders 60.3% (95% CI 54, 66.3%) | Yes |
| 2. | Badellino, et al.2021 | Argentina | 3/29/2020 - 4/12/2020 | Total sample: 2051. Final sample: 1985. Final sample without health care workers: 1659 | 18 years or older | Convenience | GAD-7<br>PHQ-9<br>PSS-10<br>PSQI | Cross-sectional | - | Anxiety symptoms<br>Mild: 38.1% (N= 632)<br>Moderate: 11.2% (N= 185)<br>Severe: 3.5% (N= 59)<br><br>Depressive symptoms<br>Mild: 22.1% (N= 366)<br>Moderate: 19.7% (N= 327)<br>Severe: 6.3% (N=105)<br><br>Stress<br>Yes: 23.6% (N=391)<br><br>Sleep quality<br>97 / 5000 | Yes |

| | Authors | Country | Data collection period | N | Inclusion criteria | Sampling | Assessment tools | Design | Time points | Results | Lockdown |
|---|---|---|---|---|---|---|---|---|---|---|---|
| 3. | Badellino, et al.2021 | Argentina | T1: 3/29/2020-4/12/2020<br>T2: 5/23/2020-6/12/2020 | T1 (N: 1985).<br>T2 (N: 2839) | 18 years or older | Convenience | PHQ-9<br>PSQI | Longitudinal | T1 and T2 during COVID-19 pandemic | Depressive symptoms<br>T1: 24.3% (N: 482)<br>T2: 47.8% (N: 135)<br><br>Conciliation insomnia<br>T1: 51.1% (N: 1023)<br>T2: 0.5 % (N: 1719)<br><br>Night awakenings<br>T1: 54.3% (N: 1078)<br>T2: 4.4 % (N: 1546)<br><br>Sleep < 7 hs<br>T1: 27.8 % (N: 551)<br>T2: 70% (N: 1987)<br><br>Poor sleep quality<br>T1: 23.1% (N: 459)<br>T2: 32.1% (N: 912) | Yes |
| 4. | Bohn, et al.2021 | Brazil | June 2020 | Total sample: 1453.<br>Final sample: 1123 | 60 years or older, enrolled in the community program | Convenience | GDS-15 | Cross-sectional | - | Depressive symptoms: 30.4% | No |
| 5. | Caballero-Domínguez, et al.2020 | Colombia | 3/30/2020 - 4/8/2020 | 700 | Residents of Colombia | Convenience | WHO-5<br>AIS | Cross-sectional | - | Depressive symptoms: 61.1% (N: 428)<br><br>Insomnia: 42% (N: 294) | Yes |
| 6. | Calegaro, et al. 2021 | Brazil | 4/22/2021 - 5/8/2021 | 3587 | 18 years or older | Snowball | AUDIT-C<br>DASS-21<br>PCL-5 | Cross-sectional | - | Stress: 58.4%<br>Anxiety: 52.3%<br>Depression: 61.2%<br>PTSD: 24.5% | - |

| | Authors | Country | Data collection period | N | Inclusion criteria | Sampling | Assessment tools | Design | Time points | Results | Lockdown |
|---|---|---|---|---|---|---|---|---|---|---|---|
| 7. | Campos, et al.2020 | Brazil | 5/18/2020 - 6/25/2020 | 13494 | 18 years or older | Convenience | DASS-21 | Cross-sectional | - | Depressive symptoms<br>Mild: 14.5%<br>Moderate: 21.8%<br>Severe: 25%<br><br>Anxiety symptoms<br>Mild: 8.5%<br>Moderate: 19.2%<br>Severe: 16.5%<br><br>Stress<br>Mild: 15.5%<br>Moderate: 16.9%<br>Severe: 18.4% | Yes |
| 8. | Caycho-Rodríguez, et al.2021 | Peru | 8/3/2020 - 8/31/2020 | 274 | - | Convenience | MHI-5<br>PHQ-2 | Cross-sectional | - | Depressive symptoms 30.7%<br>Generalized Anxiety symptoms 25.2% | Yes |
| 9. | Cerecero-Garcia, et al.2021 | Mexico | 4/20/2020 - 4/27/2020 | Total sample: 881.<br>Final sample: 595 | Men who have sex with transgender men and women. Some of them had participated in a project of Prophylaxis prior to exposure to HIV | Convenience | CESD-10 | Longitudinal | T1 and T2 during COVID-19 pandemic (Before and during lockdown). | Depressive symptoms<br>*Enrolled in ImPrEP study*<br>During lockdown: 51.9%<br>Before lockdown: 8.6%. | Yes |

| | Authors | Country | Data collection period | N | Inclusion criteria | Sampling | Assessment tools | Design | Time points | Results | Lockdown |
|---|---|---|---|---|---|---|---|---|---|---|---|
| 10. | Cheung, et al. 2021 | International, including Brazil | 3/24/2020 - 4/30/2020 | Total sample: 20,958<br>Brazilean sample: 8375 | 18 years or older | Convenience | PHQ-9 | Cross-sectional | - | Brazil (n= 8375)<br>Suicidal ideation: 1.5%<br>Not at all: 92.4% (n= 7,740)<br>Several days: 5.1% (n= 430)<br>More than half of the days: 1% (n= 83)<br>Almost every day: 1.5% (n= 122) | - |
| 11. | Côrrea, et al.2020 | Brazil | 5/16/2020 - 5/26/2020 | 213 | 18 years or older. Be a yoga practitioner with a minimum of 6 months experience | Snowball | DASS-21 | Cross-sectional | - | Stress<br>Mild: 26.3%<br>Moderate: 15%<br>Severe: 9.4%<br>Extremely severe: 2.8%<br><br>Anxiety symptoms<br>Mild: 4.2%<br>Moderate: 15.5%<br>Severe: 4.2%<br>Extremely severe 8%<br><br>Depressive symptoms<br>Mild: 10.3%<br>Moderate: 15.5%<br>Severe: 4.7%<br>Extremely severe: 3.3% | - |
| 12. | Cortés-Álvarez, et al.2020 | Mexico | 3/20/2020 - 4/5/2020 | 1105 | 18 years or older | Snowball | IESR<br>DASS-21 | Cross-sectional | - | Distress<br>Moderate to severe: 50.3%<br><br>Depressive symptoms<br>Moderate to severe: 15.7%<br><br>Anxiety symptoms<br>Moderate to severe: 22.6%<br><br>Stress<br>Moderate to severe: 19.8% | Yes |
| 13. | da Silva Júnior, et al.2021 | Brazil | 10/27/2020 - 12/11/2020 | 5879 | University students over 18 years old | Convenience | GAD-7 | Cross-sectional | - | Generalized anxiety symptoms: 44.59% | - |

| | Authors | Country | Data collection period | N | Inclusion criteria | Sampling | Assessment tools | Design | Time points | Results | Lockdown |
|---|---|---|---|---|---|---|---|---|---|---|---|
| 14. | De Boni, et al.2021 | Brazil | 20/4/20-20/5/20 | Total sample: 22785 (Brazil and Spain) Brazilian sample: 19256 | 18 years or older | Snowball | AUDIT-C | Cross-sectional | | Risky drinking: 45.6% | - |
| 15. | de Camargo, et al.2021 | Brazil | April to May 2020 | 2000 | 18 years or older | Convenience | Kessler psychological distress scale | Cross-sectional | - | Distress: 35.9% | - |
| 16. | de Souza Costaa, et al. 2021 (Pre-print) | Brazil | 5/9/2020 - 6/30/2020 | 5635 | 18 years or older | Convenience | GSI | Cross-sectional | - | High average psychological distress: 1448 (25.1)<br>Very high psychological distress: 351 (6.3) | - |
| 17. | Del Brutto, et al.2021 | Ecuador | T1: 2019 (Pre-COVID-19)<br>T2: 2020 (During COVID-19) | 2019: 673.<br>2020: 639 | Atahualpa residents over 40 years old participa-ting in the Atahualpa project. | Convenience | PSQI | Longitudinal | T1: Pre-COVID period (2019)<br>T2: during COVID-19 pandemic (2020) | Poor sleep: 49%<br><br>185 (29%) individuals at baseline, and 311 (49%) at follow-up PSQI, were “poor sleepers”. | - |
| 18. | Etchevers, et al. 2021 | Argentina | T1:3/27/20 - 3/31/20<br>T2: 5/8/2020 - 5/12/2020 | T1: 2631<br>T2: 2068 | No information | Convenience sample | SCL-27 | Longitudinal | T1 and T2 during COVID-19 pandemic | Risk of Mental Disorder<br>T1: 4.86%<br>T2: 7.2%<br><br>Sleep problems<br>T1: 73.7%<br>T2: 76.02%<br><br>Suicidal thoughts<br>T1: 4.22%<br>T2: 6.53% | Yes |
| 19. | Fernández, et al.2020 | Argentina | 4/1/2020 - 4/17/2020 | 4408 | 18 years or older | Convenience | BSI-53 | Cross-sectional | - | Phobic anxiety: 41.3%<br>Anxiety symptoms: 31.8%<br>Depressive symptoms: 27.5%<br>Distress: 27.1% | Yes |

| | Authors | Country | Data collection period | N | Inclusion criteria | Sampling | Assessment tools | Design | Time points | Results | Lockdown |
|---|---|---|---|---|---|---|---|---|---|---|---|
| 20. | Ferreira, et al. 2021 | Brazil | May and June of 2020 | Total sample: 1316. Final sample: 1130 | 18 years or older | Snowball | DASS-21 | Cross-sectional | - | Severe depressive symptoms: Total sample: 17.43% In social distance: 19.2% Not in social distancing: 13.2% | Yes |
| 21. | Feter, et al.2021 | Brazil | 6/22/2020 - 7/23/2020 | 1767 | - | Convenience | HADS | Cross-sectional | - | Anxiety symptoms: 51.3% Depressive symptoms: 35% | During the recruitment phase, up to 73.4% of state population were in the second highest level of social distancing restriction |
| 22. | Filgueiras & Stults-Kolehmainen2021 | Brazil | 3/18/2020 - 3/22/2020 | 1460 | 18 years or older | Convenience | Perceived Stress Scale-10 Filgueiras Depression Inventory, State and Trait Anxiety Inventory: state subscale | Cross-sectional | - | Acute stress: 6.9% Depressive symptoms: 4.2% Anxiety symptoms: 8,8% (8.9 women and 8.4 men) | Yes |
| 23. | García-Espinosa, et al. 2021 | Colombia | - | 1149 | University students | Convenience | GAD-7/ PHQ-9 | Cross-sectional | - | Depressive symptoms: 47.08% Anxiety symptoms: 27.06% | - |

| | Authors | Country | Data collection period | N | Inclusion criteria | Sampling | Assessment tools | Design | Time points | Results | Lockdown |
|---|---|---|---|---|---|---|---|---|---|---|---|
| 24. | González Ramírez, et al. 2020 | Mexico | 3/27/2020 - 4/2/2020 | Total sample: 4183. Final sample: 3932 | 18 years or older | Convenience | IESR | Cross-sectional | - | Postraumatic stress symptoms: 27.7% | - |
| 25. | Goularte, et al. 2021 | Brazil | 5/20/2020 - 6/14/2020 | 1996 | 18 years or older | Convenience | IESR, PROMIS Short Form v1.0 - Depression 8a), PROMIS anxiety | Cross-sectional | - | Anxiety symptoms: 81.9%<br>Depressive symptoms: 68%<br>Sleep problems: 55.3% | -. |
| 26. | Guzmán-Muñoz, et al. 2020 | Chile | 5/30/2020 - 6/6/2020 | 1082 | 18 years or older | Convenience | SF-36 | Cross-sectional | - | Mental health problems: 49.7% | - |
| 27. | Krüger-Malpartida, et al. 2020 | Peru | - | 400 | 18 years or older | Convenience | PHQ-9, GAD-7, and CPDI | Cross-sectional | - | Anxiety symptoms<br>Minimal: 49.30%<br>Mild: 40%<br>Moderate: 7.2%<br>Severe: 3.5%<br><br>Depressive symptoms<br>Minimal: 48.5%<br>Mild: 29.5%<br>Moderate: 12%<br>Severe: 10%<br><br>Peritraumatic stress<br>Mild: 35.5%<br>Severe:9.3% | No. Post - lockdown. |
| 28. | López Steinmetz, et al. 2020 | Argentina | 3/30/2020 - 5/23/2020. | 1100 | 18 years or older | Convenience | BDI-II/ STAI | Cross-sectional | - | Depressive symptoms: 29.64%<br>State anxiety: 48.55%<br>Trait anxiety: 47.91% | Yes |

| | Authors | Country | Data collection period | N | Inclusion criteria | Sampling | Assessment tools | Design | Time points | Results | Lockdown |
|---|---|---|---|---|---|---|---|---|---|---|---|
| 29. | López Steinmetz, et al.2021 | Argentina | 5/17/2020 - 6/30/2020 | 1492 | 18 years or older | Convenience | BDI/ STAI | Longitudinal | T1, T2; and T3 during COVID-19 pandemic. T1: 17th to 29th March 2020. T2: 30th March to 8th May 2020. T2: 9th May to 30th June 2020 | Depressive symptoms<br>1° sub-period (n = 866)<br>*1° measurement:* 58.66%<br>*Follow-up:* 54.62%<br><br>2° sub-period (n = 298)<br>*1° measurement:* 60.40%<br>*Follow-up:* 62.42%<br><br>3° sub-period (n = 328)<br>*1° measurement*: 55.49%<br>*Follow-up:* 57.62%<br><br>Anxiety symptoms<br>1° sub-period (n = 866)<br>*1° measurement:* 52.54%<br>*Follow-up:* 50.23%<br><br>2° sub-period (n = 298)<br>*1° measurement:* 55.03%<br>*Follow-up:* 54.36%<br><br>3° sub-period (n = 328)<br>*1° measurement:* 53.05%<br>*Follow-up: 49.08%* | Yes |

| | Authors | Country | Data collection period | N | Inclusion criteria | Sampling | Assessment tools | Design | Time points | Results | Lockdown |
|---|---|---|---|---|---|---|---|---|---|---|---|
| 30. | López-Morales, et al.2021 | Argentina | T1: 3/22-25/2020.<br>T2: 4/3-9/20<br>T3: 5/6-10/2020 | T1 and T2: 204. T3: 201 | Women over 18 years | Convenience | BDI-II<br>STAI | Longitudinal | T1, T2, and T3 during COVID-19 pandemic.<br>T1: 22-25/3/20<br>T2: 3-9/4/20<br>T3: 6-10/5/20 | Pregnant women<br>Depressive symptoms<br>T1<br>Mild: 12.7%<br>Moderate: 8.8%<br>Severe: 0%<br>T2<br>Mild: 23.5%<br>Moderate: 7.1%<br>Severe: 7.1%<br>T3<br>Mild: 20.8%<br>Moderate: 19.8%<br>Severe: 12.9%<br><br>Not pregnant women<br>Depressive symptoms<br>T1<br>Mild: 11.1%<br>Moderate: 2.0%<br>Severe: 0%<br>T2<br>Mild: 16.2%<br>Moderate: 6.1%<br>Severe: 0%<br>T3<br>Mild: 19.0%<br>Moderate: 6.0%<br>Severe: 4% | Yes |
| 31. | Loret de Mola, et al. 2021 | Brazil | T1: During 2019<br>T:2 May to July 2020 | 1136 | Brazilian mothers | Convenience | EPDS<br>GAD-7<br>IES | Longitudinal | T1: Pre-COVID period (2019)<br>T2: during COVID-19 pandemic (2020) | Stress (moderate to severe): 40.5%<br>Depressive symptoms: 29.3%<br>Generalized anxiety symptoms: 25.9% | - |

| | Authors | Country | Data collection period | N | Inclusion criteria | Sampling | Assessment tools | Design | Time points | Results | Lockdown |
|---|---|---|---|---|---|---|---|---|---|---|---|
| 32. | Martínez, et al. 2020 | Brazil | 5/11/2020 - 5/15/2020 | Total sample 2230. Final sample: 1017 | 18 years or older | Convenience | HADS | Cross-sectional | - | Depressive symptoms: 48.8%<br>Anxiety symptoms: 82.6% | Yes |
| 33. | Meda-Lara, et al. 2021 | Mexico | 4/2/2020 - 5/4/2020 | 666 | - | Convenience | DASS-21<br>ESR | Cross-sectional | - | Depressive symptoms<br>Mild: 8.71%<br>Moderate: 6.61%<br>Severe: 3.45%<br>Extremely severe: 3.6%<br><br>Anxiety symptoms<br>Mild: 5.4%<br>Moderate: 5.86%<br>Severe: 3.3%<br>Extremely severe: 6.16%<br><br>Stress<br>Mild: 16.22%<br>Moderate: 6.31%<br>Severe: 3.6%<br>Extremely severe: 1.8% | - |
| 34. | Miranda, et al. 2021 | Argentina | May to July 2020 | 305 | Women over 18 years of age in the postpartum period | Convenience | ISI<br>PDSS-SF | Cross-sectional | - | Depressive symptoms: 37%<br>Insomnia: 46% | Yes |
| 35. | Montero Doig, et al.2021 | Peru | T1: Pre-pandemic period<br>T2: Pandemic period<br>Unspecified dates | 150 | Adult population of the south east of Madre de Dios | Convenience | Anxiety state (A/S) and Anxiety trait (A/T) | Longitudinal | T1: Pre-COVID period (2019)<br>T2: during COVID-19 pandemic (2020) | State anxiety: 65%.<br>Before COVID-19<br>Mild anxiety: 26.67%<br>Moderate anxiety: 46.67%<br>Severe anxiety: 26.67%<br>During COVID-19<br>Mild anxiety: around 0%<br>Moderate anxiety: 53.33%<br>Severe anxiety: 46.67% | Yes |

| | Authors | Country | Data collection period | N | Inclusion criteria | Sampling | Assessment tools | Design | Time points | Results | Lockdown |
|---|---|---|---|---|---|---|---|---|---|---|---|
| 36. | Monterrosa-Blanco, et al.2021 | Colombia | 5/1/2020 - 5/30/2020 | 984 | Women between 40 to 59 years | Convenience | Jong Gierveld Loneliness Scale | Cross-sectional | - | Emotional loneliness 44% (n= 433)<br>Social loneliness 42.2% (n= 415)<br>General loneliness 44.5% (n= 438) | Yes |
| 37. | Monterrosa-Castro, et al.2021 | Colombia | 6/1/2020 - 6/5/2020 | Total sample 1185. Final sample: 1133 | Women aged between 40-79 years | Convenience | DJGLS<br>MRS<br>FCV-19S-5<br>CAS<br>Francis-5 Religiosity Scale | Cross-sectional | - | Emotional loneliness: 43.1% (n=489)<br>Social loneliness: 39.9% (n=452)<br>General loneliness: 43.3% (n=491) | Yes |
| 38. | Naranjo-Hernández, et al.2021 | Cuba | May to June 2020 | 100 | Functional older adults | Convenience | HAD<br>EEP-10-C | Cross-sectional | - | Mild anxiety: 73% (n=73)<br>Severe anxiety: 21% (n=21)<br><br>Mild depression: 50% (n=50)<br>Severe depression: 24% (n=24) | Yes |
| 39. | Nomura, et al.2021 | Brazil | 6/1/2020 - 8/31/2020 | Total sample: 1683. Final sample: 1662 | Maternal age more than 18 years; gestational age more than 36 weeks at childbirth; single alive newborn without malformations; and absence of mental disorders | Convenience | BAI | Cross-sectional | - | Mild anxiety: 22.4% (n=372)<br>Moderate anxiety: 13.9% (n=231)<br>Severe anxiety: 9.6% (n=159) | Yes |
| 40. | Oblitas Gonzales, et al.2020 | Peru | 3/16/2020 - 6/30/2020 | 67 | 18 years or older | Convenience | EAA | Cross-sectional | - | Mild anxiety: 20.9% (n=14)<br>Moderate anxiety: 13.4% (n=9)<br>Severe anxiety: 9% (n=6) | Yes |

| | Authors | Country | Data collection period | N | Inclusion criteria | Sampling | Assessment tools | Design | Time points | Results | Lockdown |
|---|---|---|---|---|---|---|---|---|---|---|---|
| 41. | Paico, et al.2021 | Peru | - | 203 | Post graduate student | Convenience | STAI<br>AQ | Cross-sectional | - | (Sate-Anxiety%)<br>Above average: 15.3%<br>Elevated: 84.7%<br><br>(Trait-Anxiety%)<br>Above average: 44.3%<br>Elevated: 9.9% | Yes |
| 42. | Passos, et al.2020 | Brazil and Portugal | 5/27/2020 - 7/8/2020 | Total sample: 550. Brazilian sample: 289 | 18 years or older | Snowball | SWLS<br>GAD-7<br>PHQ-2 | Cross-sectional | - | *-Brazil-*<br><br>Anxiety: 74.7% *(*n=216*)*<br>Depression: 26.6% *(*n=77*)* | Variation between regions. |
| 43. | Pereira-Ávila, et al.2021 | Brazil | 4/17/2020 - 5/15/2020 | 900 | Individuals aged 60 or over were adopted as inclusion criteria, and foreigners residing in Brazil were excluded | Convenience | PHQ-9 | Cross-sectional | - | Depressive symptoms:<br>91,9% mild depressive symptoms<br>2% moderately severe symptoms<br>1,4% symptoms of severe depression | Yes |
| 44. | Pérez-Cano, et al.2020 | Mexico | 3/22/2020 - 3/30/2020 | 613 | 18 years or older | Convenience | DASS-21<br>STAI | Cross-sectional | - | Depression: 41.3%<br>Moderate/Severe Anxiety: 48.8%<br>State-Anxiety: 42% | - |
| 45. | Porter, et al.2021 | Peru<br>Ethiopia<br>India<br>Vietnam | August to October 2020 | Total sample: 9730. Final sample: 8988. **Peruvian sample**: 1887 | 18 years or older | Convenience | PHQ-8<br>GAD-7 | Cross-sectional | - | *-Peru-*<br><br>Mild depression: 32%<br>Moderate/severe depression: 9.6%<br><br>Mild anxiety: 41%<br>Moderate/severe anxiety: 13.5% | - |

| | Authors | Country | Data collection period | N | Inclusion criteria | Sampling | Assessment tools | Design | Time points | Results | Lockdown |
|---|---|---|---|---|---|---|---|---|---|---|---|
| 46. | Puccinelli, et al.2021 | Brazil | 6/2/2020 - 6/12/2020 | Total sample: 2140. Final sample: 1853 | 18 years or older | Convenience | IPAQ<br>PHQ-9<br>GAD-7 | Cross-sectional | - | Mild depression: 36.2% (n= 670)<br>Moderate depression: 16.4% (n= 304)<br>Moderate-Severe depression: 8,4% (n= 155)<br>Severe depression: 4.8% (n= 89)<br><br>Mild anxiety: 36.4% (n=674)<br>Moderate anxiety: 14.3% (n=265)<br>Severe anxiety: 8.9% (n=164) | No |
| 47. | Puccinelli, et al.2021 | Brazil and Switzerland | 6/2/2020 - 6/12/2020 | Total sample: 114. Brazilian sample: 57. | 18 years or older | Convenience | IPAQ<br>PHQ-9<br>GAD-7 | Cross-sectional | - | Mild depression: 29.8% (45.6% Brazil, 14% Suiza)<br>Moderate/Severe depression: 14.9% (22.8% Brazil, 7% Suiza)<br><br>Mild anxiety: 33.3% (47.4% Brazil, 19.3% Suiza)<br>Moderate/severe anxiety: 10.5% (17.5% Brazil, 3.5% Suiza) | No |
| 48. | Quiroga-Garza, et al.2021 | Mexico | 4/30/2020 - 6/16/2020 | 604 | 18 years or older | Convenience | DASS-21<br>PERMA Profiler<br>Brief COPE<br>MCCS | Cross-sectional | - | Moderate-severe psychological distress: 29.5%<br>Extremely severe psychological distress: 16.4% | Yes |
| 49. | Ramírez-Coronel, et al.2021 | Ecuador | - | 381 | Adult women between 18 and 65 years of age who were in confine-ment | Convenience | STAI<br>IDER | Cross-sectional | - | Anxiety: 71.4% (n=272)<br>Depression: 77.2% (n=294) | - |

| | Authors | Country | Data collection period | N | Inclusion criteria | Sampling | Assessment tools | Design | Time points | Results | Lockdown |
|---|---|---|---|---|---|---|---|---|---|---|---|
| 50. | Ramos-Padilla, et al.2021 | Ecuador | June to July 2020 | 9522 | 18 years or older | Convenience | PSQI | Cross-sectional | - | Sleep quality (bad quality):<br>Women: 51%<br>Men: 47%<br><br>Sleep duration (<7 hours)<br>Women: 34.9%<br>Men: 36.8%<br><br>Sleep efficiency (<65%)<br>Women: 23.6%,<br>Men: 23.7% | Yes |
| 51. | Ribeiro, et al.2021 | Brazil | 4/27/2020 - 7/5/2020 | Total sample: 981. Final sample: 936 | Adults over 18 years of age who are residents of cities with established social distance restrictions | Snowball | GAD-7<br>CES-D | Cross-sectional | - | Severe anxiety: 17.36% (n=162)<br>Severe depression: 66.13% (n= 617) | - |
| 52. | Riter, et al.2021 | Brazil | 5/27/2020 - 6/15/2020 | 232 | Parents over 18 years of age | Convenience | SRQ-20<br>PSS-4<br>PSOC | Cross-sectional | - | Stress: 92.2% above the cut-off point.<br>Common mental disorders:46.6% above the cut-off point. | - |
| 53. | Ruiz-Frutos, et al.2021 | Peru | 4/2/2020 - 9/2/2020 | 1699 | 18 years or older | Snowball | GHQ-12 | Cross-sectional | - | Pychological distress: 59.7% (n= 1014)<br>Men: 64.9%<br>Women: 53%<br>40 years old or less: 65.7% ; 53.4% in ≥40 | - |
| 54. | Schmitt, et al.2021 | Brazil | 4/14/2020 - 4/23/2020 | 3274 | 18 years or older | Snowball | PHQ-9<br>EUROHIS-QOL<br>MOS<br>WHOQoL-SRPB<br>CD-RISC | Cross-sectional | - | Depression: 16.13% (n= 528)<br><br>Clarification: the exclusion of the N of healthcare practitioners with depression was calculated (n = 247)<br>Depression % en the total sample: 23.67% (n= 775) | - |

| | Authors | Country | Data collection period | N | Inclusion criteria | Sampling | Assessment tools | Design | Time points | Results | Lockdown |
|---|---|---|---|---|---|---|---|---|---|---|---|
| 55. | Scotta, et al.2020 | Argentina | March 2020 | 584 | University students | Convenience | ISI<br>PSWQ<br>CRI<br>UWES-S | Cross-sectional | - | Moderate insomnia: 23%<br>Severe insomnia: 4% | Yes |
| 56. | Seco Ferreira, et al.2020 | Brazil | 4/3/2020 - 4/16/2020 | 924 | - | Convenience | DASS-21<br>IUS-12 | Cross-sectional | - | Anxiety: 48.5%<br>Depression: 41.9%<br>Stress: 44.8% | Yes |
| 57. | Serafim, et al.2021 | Brazil | 5/22/2020 - 6/5/2020 | Total sample: 3031. Final sample: 3000 | 18 years or older | Snowball | DASS-21 | Cross-sectional | - | Anxiety: 39.7% (n=1191)<br>Mild 18.3% (n= 218)<br>Moderate 41.4% (n= 493)<br>Severe40.3% (n= 480)<br>Depression: 46.4% (n= 1392)<br>Mild 32.5% (n= 453)<br>Moderate 35.1% (n= 489)<br>Severe 32.1% (n= 450)<br>Stress: 42.2% (n= 1266)<br>Mild 31.7% (n= 401)<br>Moderate 33.3% (n= 422)<br>Severe 35% (n= 443) | - |
| 58. | Silva, et al.2020 | Brazil | 5/12/2020 - 5/14/2020 | 1154 | - | - | DASS-21 | Cross-sectional | - | Moderate-extremely severe anxiety: women 46.41% (n= 374); men 28.74% (n= 100)<br>Moderate-extremely severe depression: women 60.43% (n= 487); men 40.81% (n= 142)<br>Moderate-extremely severe stress: women 51.74% (n= 417): men 31.61% (n= 110) | - |

| | Authors | Country | Data collection period | N | Inclusion criteria | Sampling | Assessment tools | Design | Time points | Results | Lockdown |
|---|---|---|---|---|---|---|---|---|---|---|---|
| 59. | Soares, et al.2021 | Brazil | 8/1/2020 - 9/1/2020 | 206 | Athletes | Convenience | STAI<br>BRUMS | Cross-sectional | - | Trait anxiety<br>Moderate: 72.12% (n= 154)<br>Elevated: 24.87% (n= 51)<br>State anxiety<br>Moderate: 71.21% (n= 146)<br>Elevated: 28.78% (n= 59)<br>Sleep Quality<br>Too bad: 5.78% (n=7)<br>Bad: 15.7% (n=27) | - |
| 60. | Souza, et al.2021 | Brazil | April to May 2020 | Total sample: 3793. Final sample: 3200 | 18 years or older | Snowball | DASS-21 | Cross-sectional | - | Anxiety: 19.4%<br>Mild: 7.9% (n= 252)<br>Moderate: 17.3% (n= 552)<br>Severe: 6.3% (n=203)<br>Extreme: 13.1% (n= 419)<br>Depression: 21.5%<br>Mild: 12.9% (n= 414)<br>Moderate: 18.8% (n= 602)<br>Severe: 6.3% (n= 203)<br>Extreme: 13.1% (n= 419)<br>Stress: 21.5%<br>Mild: 12.9% (n= 414)<br>Moderate: 13.1% (n= 419)<br>Severe: 12.9% (n= 413)<br>Extreme: 8.6% (n= 275) | - |
| 61. | Stults-Kolehmainen, et al.2021 (Pre-print) | Brazil | T1: 3/20/2020 - 3/25/2020<br>T2: 15/4/20 - 20/4/20 | 360 | - | Convenience | PSS-10<br>FDI<br>STAI-S | Longitudinal | T1 and T2 during COVID-19 | Perceived stress: 237 (65.8%) and 269 (74.7%) of participants scored above the population mean at time 1 and 2, respectively.<br>Excessive stress: 6.9% (IC 95 5.2%-8.6%) in the first round | Yes |

| | Authors | Country | Data collection period | N | Inclusion criteria | Sampling | Assessment tools | Design | Time points | Results | Lockdown |
|---|---|---|---|---|---|---|---|---|---|---|---|
| | | | | | | | | | | and 9.7% (IC 95 8.2%-11.2%) in the second round. High depression: 4.2% (IC 95 3.6%-4.8%) at time 1 and 8.0% (IC 95 7.1%-8.9%) at time 2. Excessive state anxiety: 8.7% (IC 95 7.4%-10.0%) in the first round, 14.9% (IC 95 12.3%-17.5%) in the second round. | |
| 62. | Suárez-Rico, et al.2021 | Mexico | August to September 2020 | 293 | New mothers over 18 years of age 4 to 12 weeks postpartum. | Convenience | EPDS STAI PSS-10 | Cross-sectional | - | State anxiety: 46.1% (n= 135) Depression: 39.2% (n= 115) Low stress: 31.1% (n= 91) Moderate stress: 58% (n= 170) Elevated stress: 10.9% (n= 32) | Yes |
| 63. | Terán-Pérez, et al.2021 | Mexico | 3/28/2020 - 5/26/2020 | 1230 | 18 years or older | Convenience | PSQI GAD-7 PHQ-9 | Cross-sectional | - | Mild anxiety: women 34.8%; men 38.1% Moderate anxiety: women 18.3%; men 16.9% Severe anxiety: women 18%; men 19%. Moderate and severe depression: women 24.5%; men 18.6% Poor sleep quality: women 79%; men 60% | Yes |

| | Authors | Country | Data collection period | N | Inclusion criteria | Sampling | Assessment tools | Design | Time points | Results | Lockdown |
|---|---|---|---|---|---|---|---|---|---|---|---|
| 64. | Toledo-Fernández, et al.2021 | Mexico | T1: 4/8/2020 - 4/18/2020<br>T2: 5/11/2020 - 5/27/2020 | Total sample: 670. Final sample: 552 | - | Convenience | IES-6<br>PHQ-9<br>GAD-7 | Longitudinal | T1 and T2 during COVID-19 pandemic. | Moderate-severe anxiety<br>T1: 11.94%<br>T2: 12.23%<br><br>Moderate-severe depression<br>T1: 5.22%<br>T2: 6.26%<br><br>Psychological distress<br>T1: 27.61%<br>T2: 21.94% | - |
| 65. | Torales, et al.2020 | Paraguay | 10/10/2020 - 4/15/2020 | 2206 | 18 years or older | Convenience | PSS-10 | Cross-sectional | - | (Stress%)<br>*Total sample:*<br>-Moderate stress: 67,95% (n= 1499)<br>High stress: 9.84% (n= 217)<br>*Participants without mental disorder*<br>- Moderate stress 67.49% (n= 1304)<br>- High stress 8.02% (n= 155)<br>*Participants with mental disorder*<br>- Moderate stress: 71.17% (n= 195)<br>- High stress: 22.63% (n= 62) | Yes |
| 66. | Torrente, et al.2021 | Argentina | 3/24/2020 | 10053 | 18 years or older | Convenience | PHQ-9<br>GAD-7 | Cross-sectional | - | Anxiety: 23.2%<br>Mild: 31.6%<br>Moderate: 13.6%<br>Severe: 8.1%<br><br>Depression: 33.7%<br>Mild: 18.5%<br>Moderate: 18.1%<br>Severe: 10.5% | Yes |

| | Authors | Country | Data collection period | N | Inclusion criteria | Sampling | Assessment tools | Design | Time points | Results | Lockdown |
|---|---|---|---|---|---|---|---|---|---|---|---|
| 67. | Torrente, et al. 2021 | Argentina | 5/21/2020 and enabled for 15 days (6/5/2020) | 3617 | 18 years or older | Convenience sample | PHQ-9<br>GAD-7<br>UCLA-LS | Cross-sectional | - | Depression<br>45.6%<br>Mild 24.5%<br>Moderate 22.3%<br>Severe 15.4%<br><br>Anxiety<br>27%<br>Mild 26.3%<br>Moderate 16.5%<br>Severe 10.5%<br><br>Loneliness<br>Low 66.3%<br>Medium 17.1%<br>High 10.4%<br>Extreme: 6.2% | Yes |
| 68. | Tyler, et al.2021 | 15 countries of Latin America and the Caribbean | 4/19/2020 - 5/3/2020 | Total sample: 823.<br>Latin American and Caribbean sample: 366 | 60 years or older | Snowball | DASS-21<br>GAD-7<br>EPII | Cross-sectional | - | Depressive symptoms: 10.35%<br>Anxiety symptoms: 5.98% | - |
| 69. | Vitorino, et al. 2021 (Pre-print) | Brazil | 1/5/2020 - 3/6/2020 | Total sample: 1167. Final sample: 1156 | Adults over 18 years of age with at least 15 days in isolation | Convenience | PHQ-9<br>GAD-7<br>WHOQOL-BREF | Cross-sectional | - | Depression: 41.9%<br>Anxiety: 29% | Yes |
| 70. | Vizioli & Crespi2021 | Argentina | 3/20/2020 - 4/20/2020 | 430 | 18 years or older | Convenience | SA-45 | Cross-sectional | - | Anxiety symptoms: 15%<br>Depressive symptoms: 14% | Yes |
| 71. | Zhang, et al.2021 | Brazil | 3/25/2020 - 3/28/2020 | 638 | 18 years or older | Convenience | CPDI | Cross-sectional | - | Mild-moderate distress: 52% (n= 332)<br>Severe distress: 18.8% (n= 120) | Yes |

#### Tables S2 – S4. Regression analyses

*Table S2. Regression coefficients for predicting depression symptoms*

| Variable | B | SE | β | t | p |
|---|---|---|---|---|---|
| Cases (1/millon) | -1.16 | .95 | -.67 | -1.23 | .227 |
| Deaths (1/millon) | 6.76 | 9.99 | .33 | .68 | .502 |
| Lockdown stringency index | .15 | .29 | .08 | .50 | .617 |
| Days since beginning of the pandemic | .14 | .09 | .34 | 1.60 | .117 |

*Note*. $R^2_{adj}$ = -.005 (N = 44, p = .447)

*Table S3. Regression coefficients for predicting anxiety symptoms*

| Variable | B | SE | β | t | p |
|---|---|---|---|---|---|
| Cases (1/millon) | 4.33 | 3.04 | 2.06 | 1,43 | .164 |
| Deaths (1/millon) | -61.85 | 38.58 | -2.22 | -1.60 | .119 |
| Lockdown stringency index | -.49 | .41 | -.21 | -1.18 | .246 |
| Days since beginning of the pandemic | .03 | .09 | .08 | .32 | .750 |

*Note*. $R^2_{adj}$ = .118 (N = 36, p = .095)

*Table S4. Regression coefficients for predicting stress symptoms*

| Variable | B | SE | β | t | p |
|---|---|---|---|---|---|
| Cases (1/millon) | .49 | 11.52 | .03 | .04 | .967 |
| Deaths (1/millon) | -111.92 | 452.25 | -.30 | -.25 | .809 |
| Lockdown stringency index | .67 | 1.88 | .11 | .36 | .728 |
| Days since beginning of the pandemic | .32 | .56 | .52 | .58 | .574 |

*Note*. $R^2_{adj}$ = -.226 (N = 17, p = .896)

#### Figure S1. Prevalence of depressive symptoms in cross-sectional studies

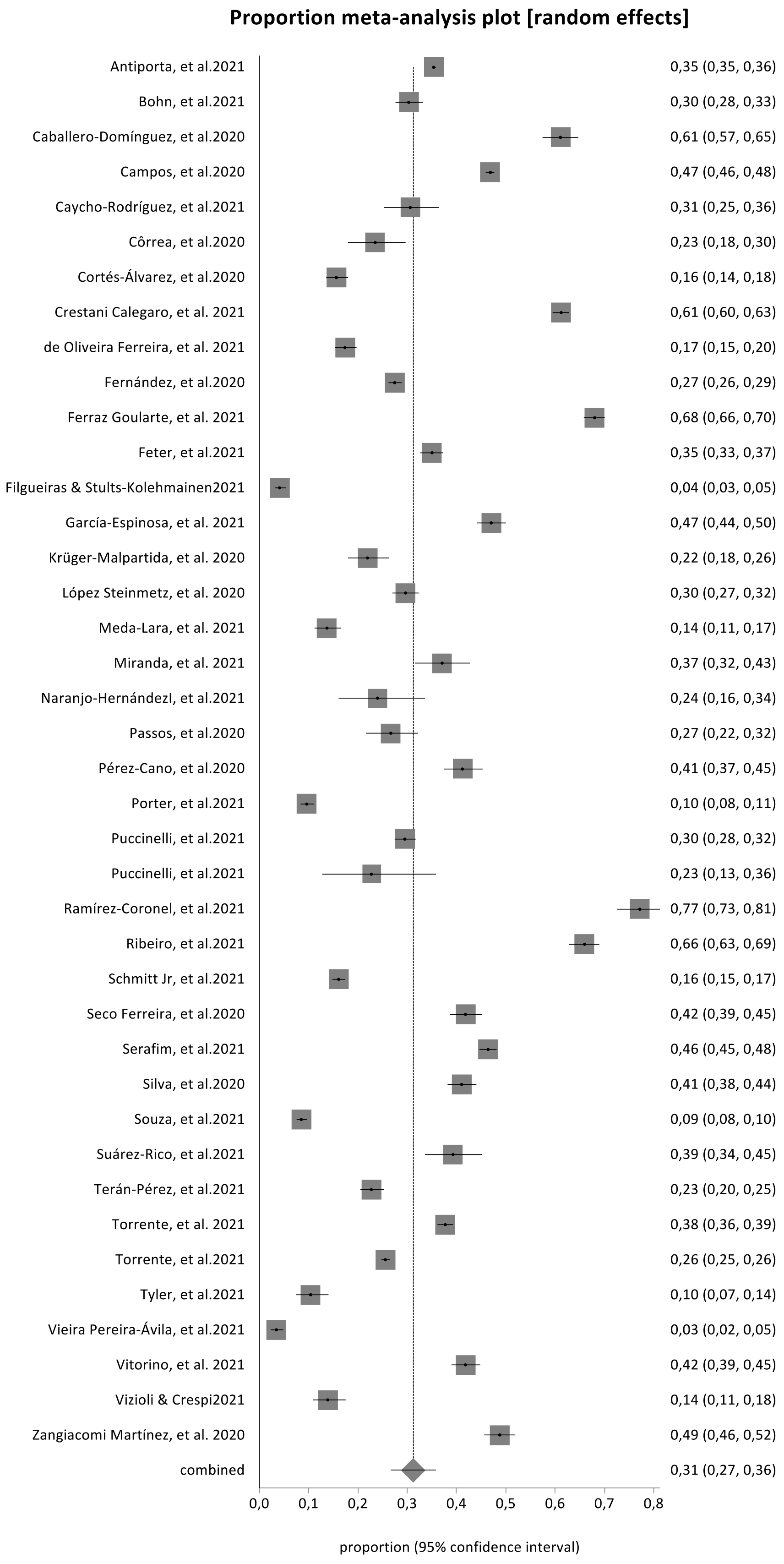

#### Figure S2. Prevalence of depressive symptoms by country

##### a. Brazil

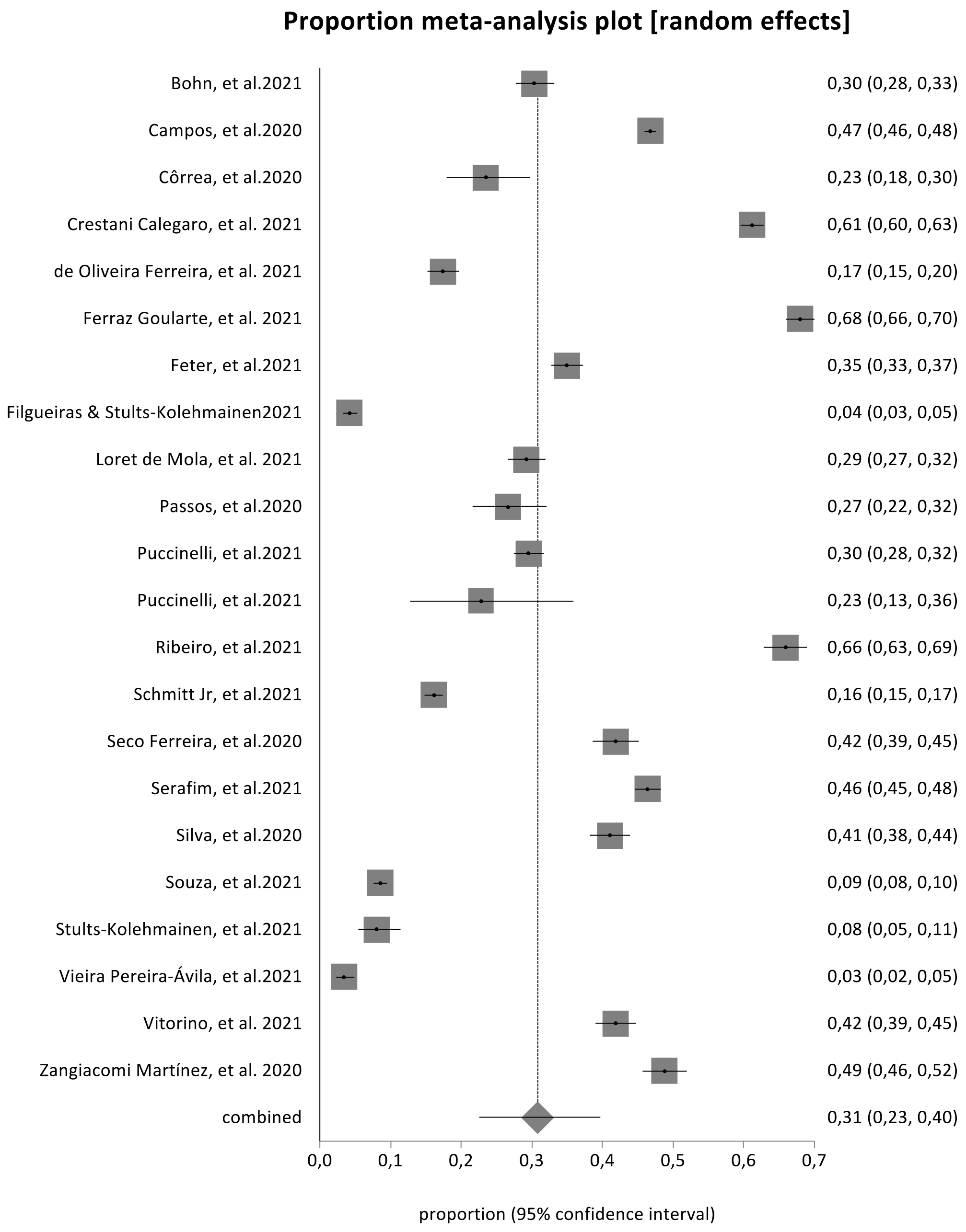

#### b. Argentina

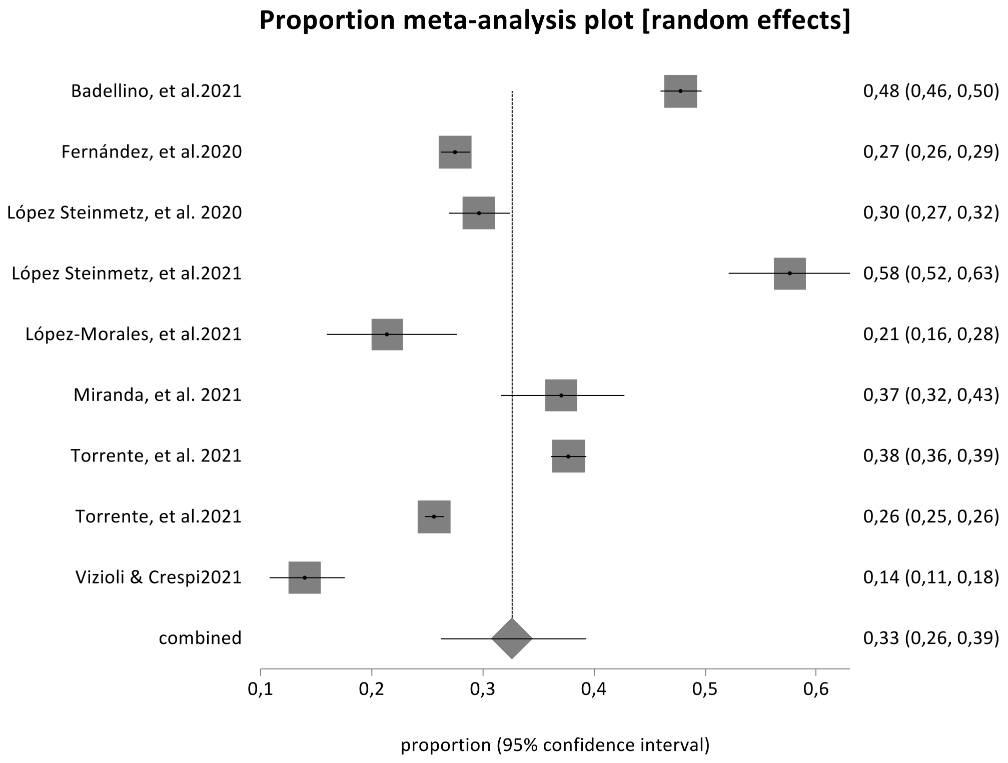


#### c. Mexico

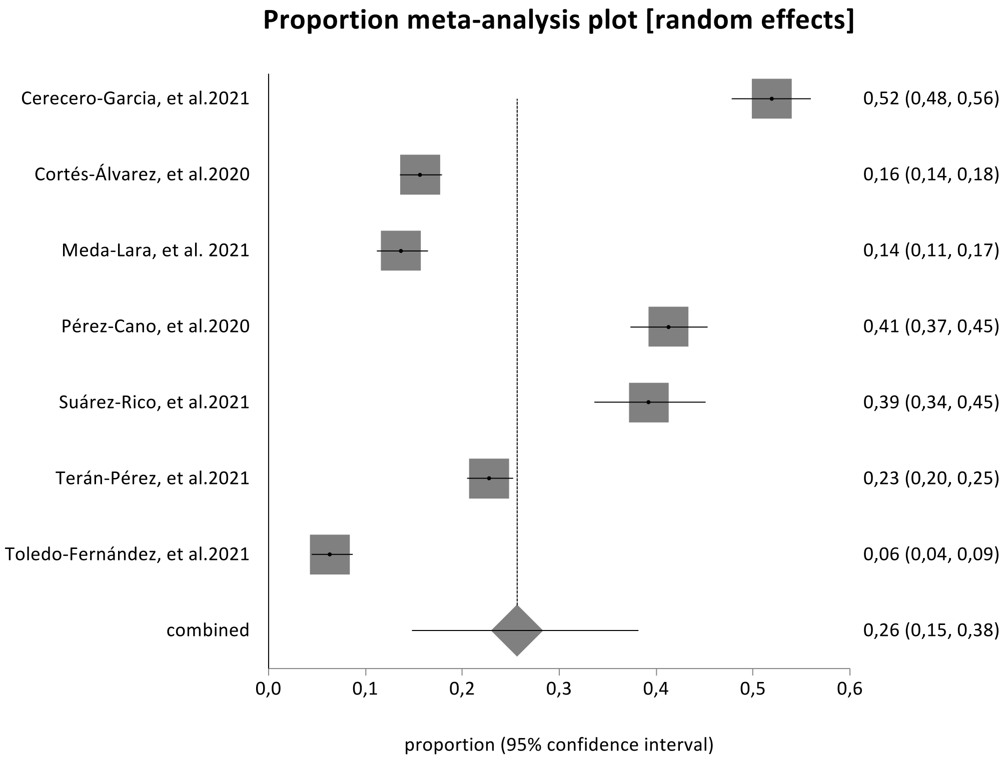

#### d. Peru

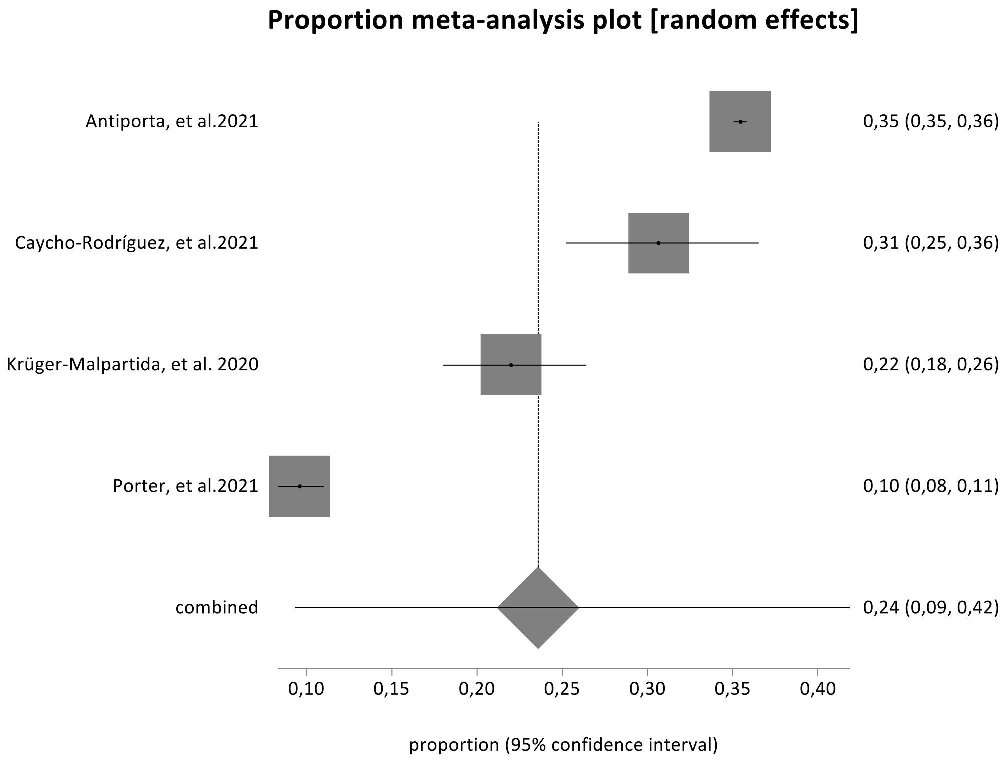

Proportion meta-analysis plot [random effects]
Antiporta, et al.2021
Caycho-Rodríguez, et al.2021
Krüger-Malpartida, et al. 2020
Porter, et al.2021
combined
0,35 (0,35, 0,36)
0,31 (0,25, 0,36)
0,22 (0,18, 0,26)
0,10 (0,08, 0,11)
0,24 (0,09, 0,42)
0,10
0,15
0,20
0,25
0,30
0,35
0,40
proportion (95% confidence interval)

### Figure S3. Prevalence of anxiety symptoms in cross-sectional studies

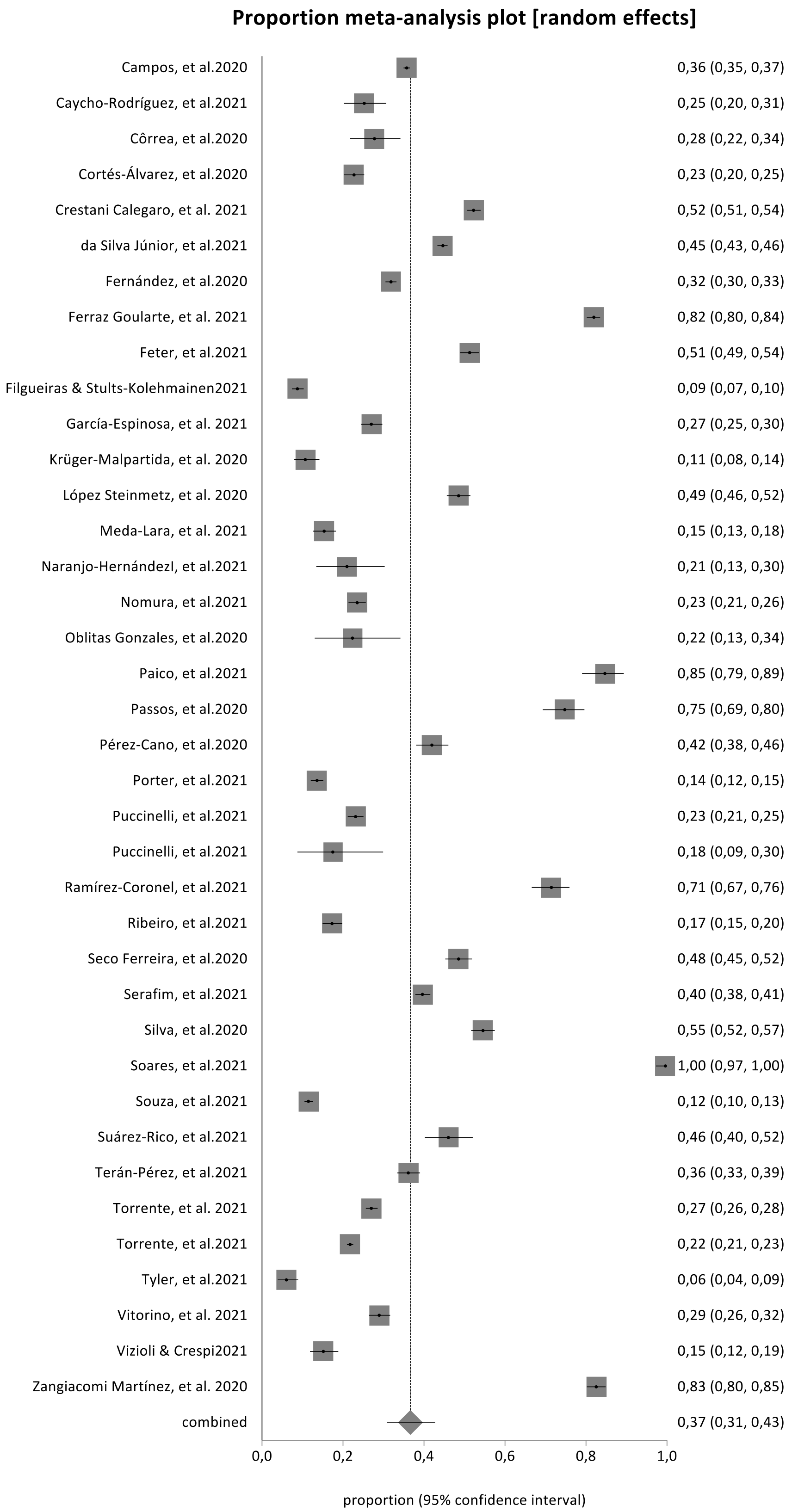

### Figure S4. Prevalence of anxiety symptoms by country

#### a. Brazil

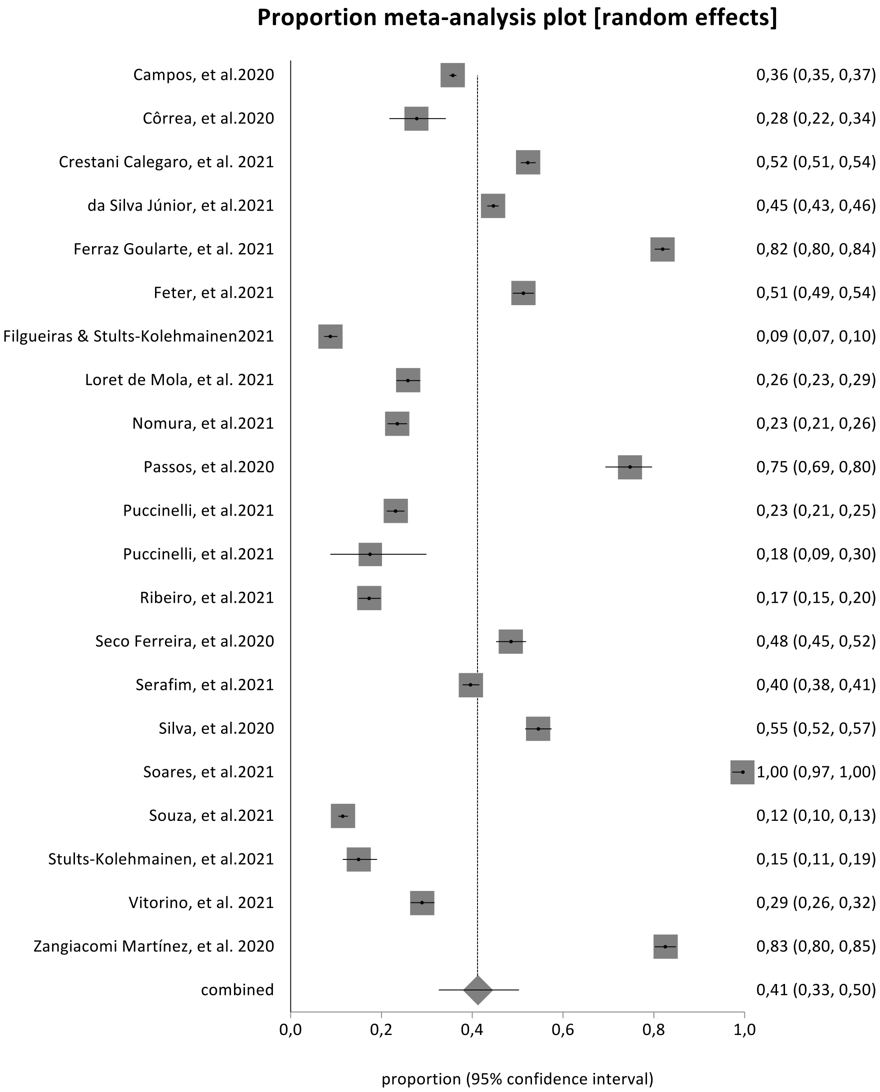

#### b. Argentina

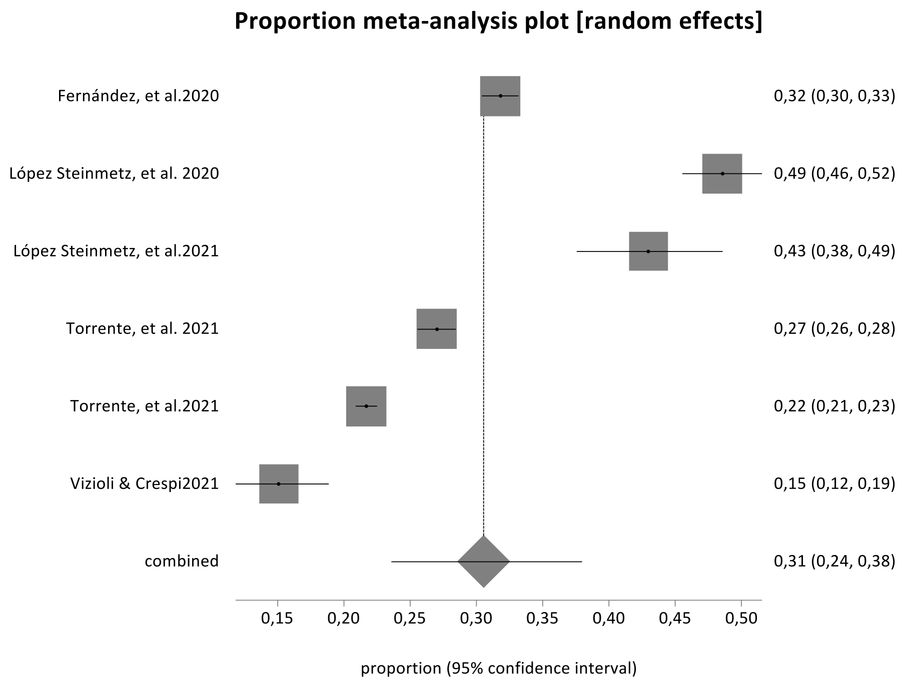


#### c. Mexico

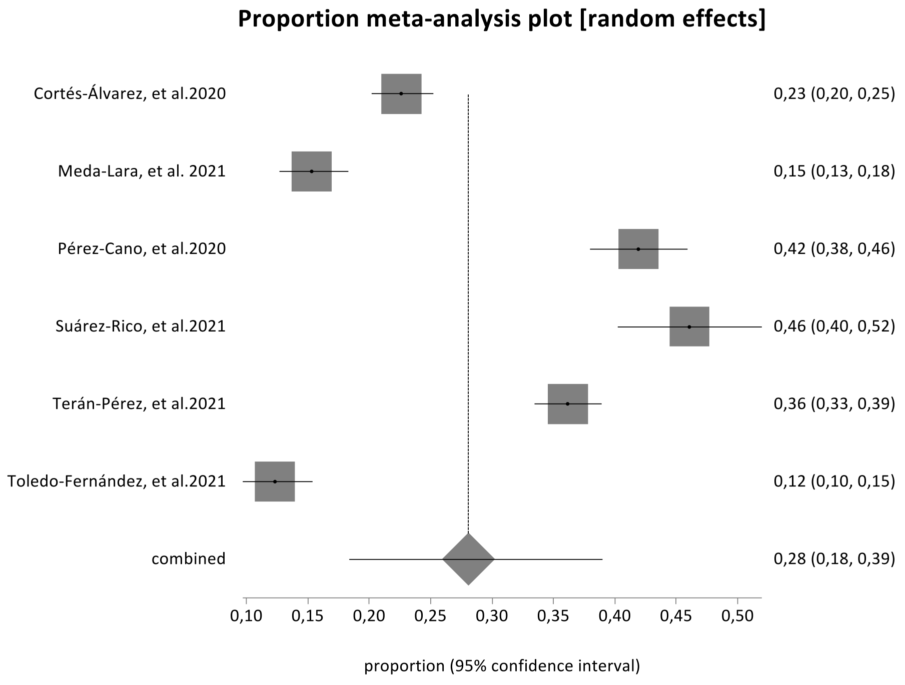

#### d. Peru

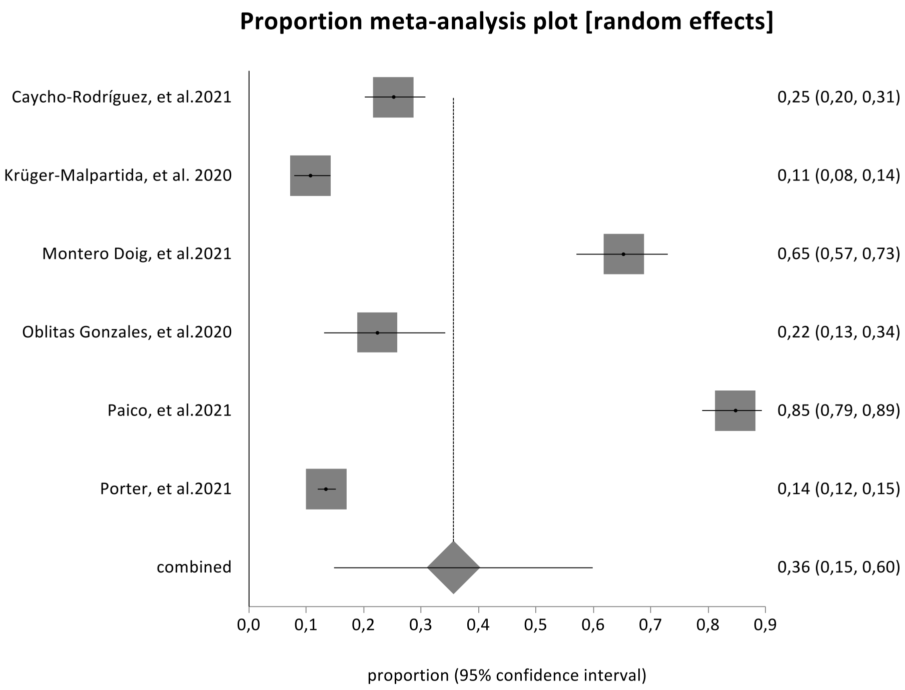

Proportion meta-analysis plot [random effects]
Caycho-Rodríguez, et al.2021
Krüger-Malpartida, et al. 2020
Montero Doig, et al.2021
Oblitas Gonzales, et al.2020
Paico, et al.2021
Porter, et al.2021
combined
0,25 (0,20, 0,31)
0,11 (0,08, 0,14)
0,65 (0,57, 0,73)
0,22 (0,13, 0,34)
0,85 (0,79, 0,89)
0,14 (0,12, 0,15)
0,36 (0,15, 0,60)
0,0 0,1 0,2 0,3 0,4 0,5 0,6 0,7 0,8 0,9
proportion (95% confidence interval)

### Figure S5: Prevalence of stress symptoms

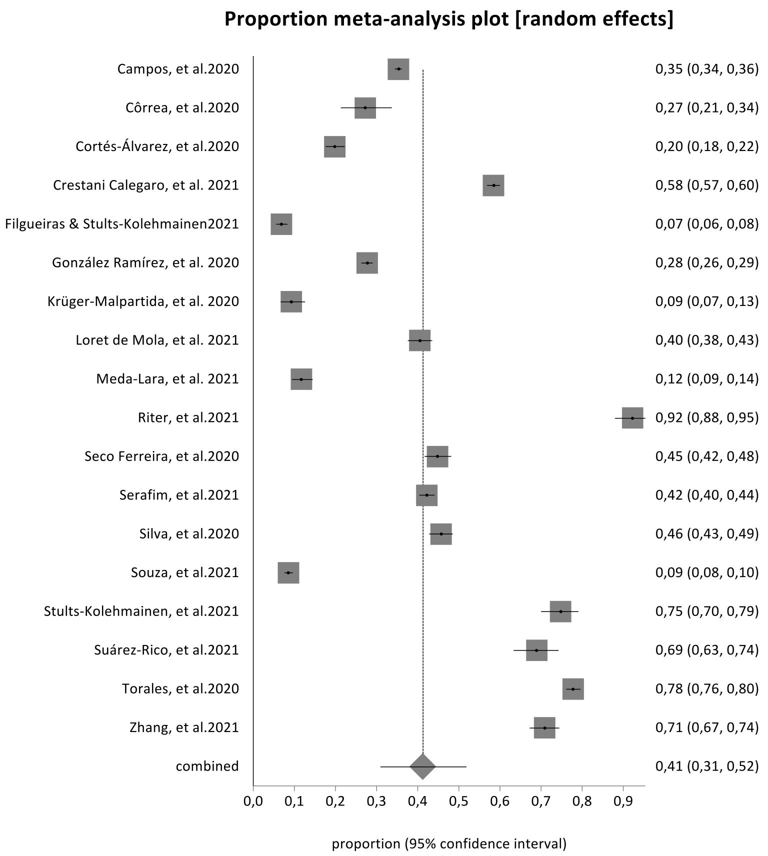


### Figure S6. Prevalence of distress symptoms

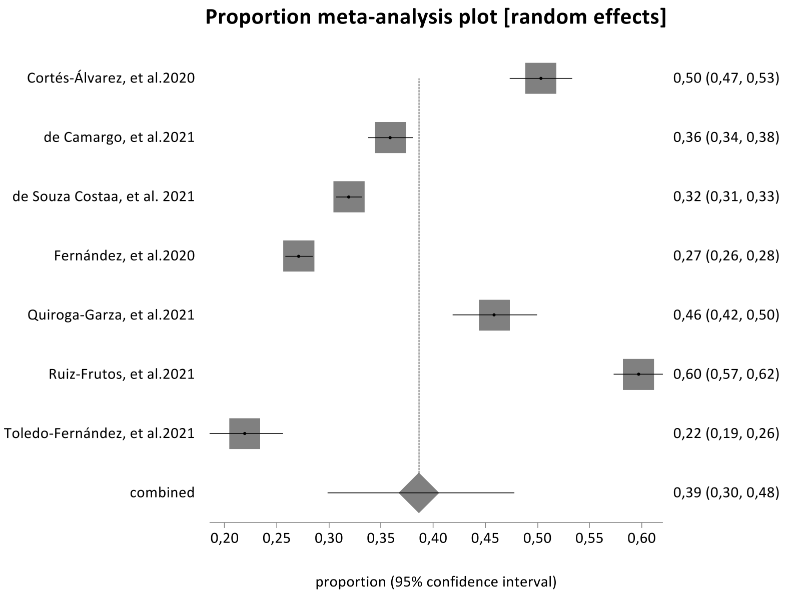

#### Figure S7. Prevalence of sleep quality problems

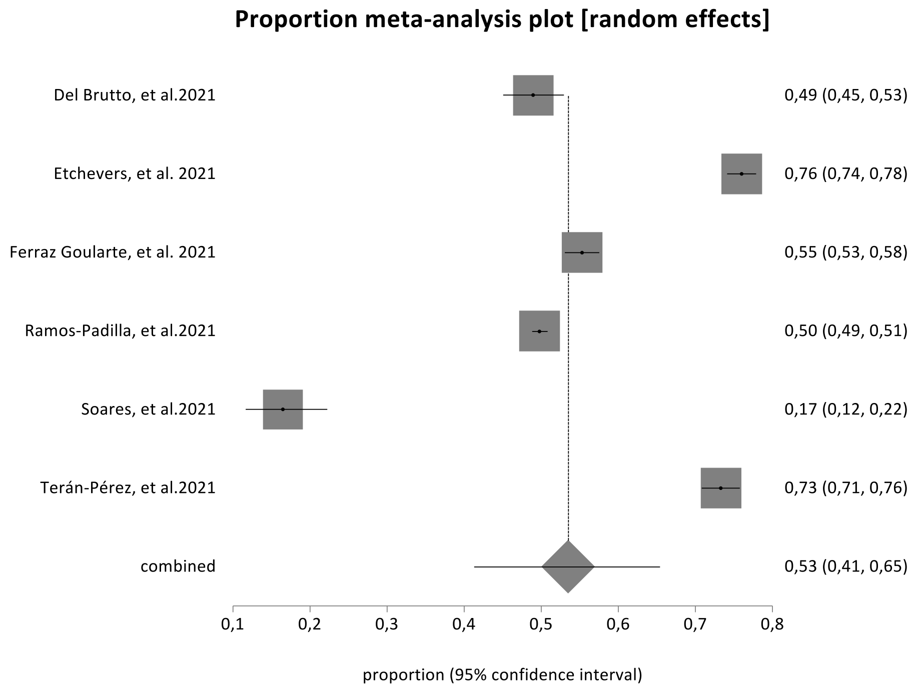


#### Figure S8. Prevalence of loneliness

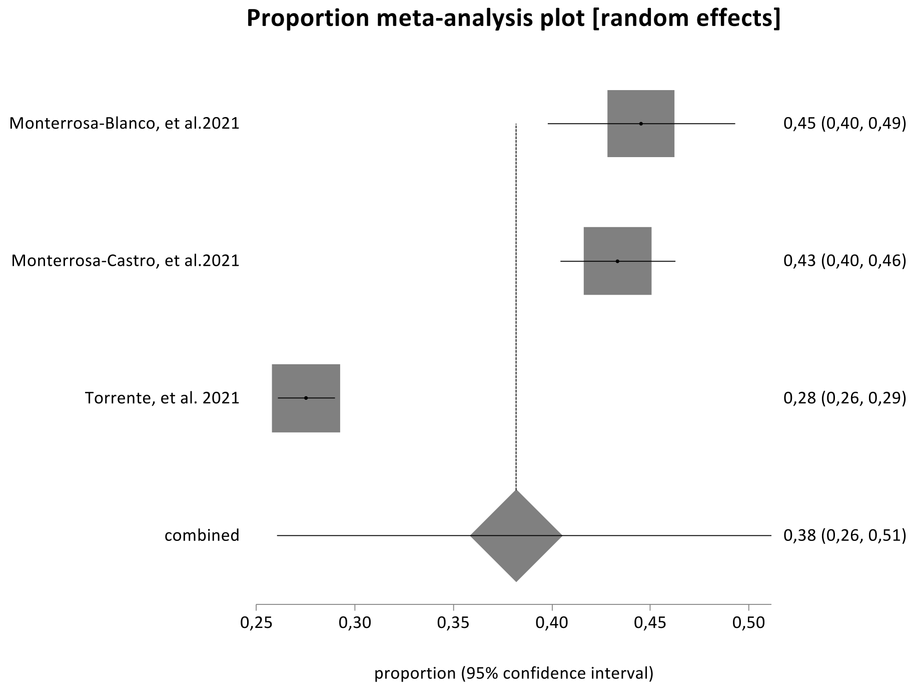

associated factors during the COVID-19 pandemic in Yoga practitioners^ien. *Rev. bras. ativ. fís. saúde*, *25*, 1-7.

22. Filgueiras, A., & Stults-Kolehmainen, M. (2021). Risk Factors for Potential Mental Illness Among Brazilians in Quarantine Due To COVID-19.

*Psychological reports*, 33294120976628. https://doi.org/10.1177/0033294120976628

23. García-Espinosa, P., Ortiz-Jiménez, X., Botello-Hernández, E., Aguayo-Samaniego, R., Leija-Herrera, J., & Góngora-Rivera, F. (2021). Psychosocial impact on health-related and non-health related university students during the COVID-19 pandemic. Results of an electronic survey. *Rev Colomb Psiquiatr*. https://doi.org/10.1016/j.rcp.2021.04.008
24. González Ramírez, L. P., Martínez Arriaga, R. J., Hernández-Gonzalez, M. A., & De la Roca-Chiapas, J. M. (2020). Psychological Distress and Signs of Post-Traumatic Stress in Response to the COVID-19 Health Emergency in a Mexican Sample. *Psychol Res Behav Manag*, *13*, 589-597. https://doi.org/10.2147/prbm.S259563
25. Goularte, J. F., Serafim, S. D., Colombo, R., Hogg, B., Caldieraro, M. A., & Rosa, A. R. (2021). COVID-19 and mental health in Brazil: Psychiatric symptoms in the general population. *Journal of Psychiatric Research*, *132*, 32-37. https://doi.org/10.1016/j.jpsychires.2020.09.021
26. Guzmán-Muñoz, E., Concha-Cisternas, Y., Oñate-Barahona, A., Lira-Cea, C., Cigarroa-Cuevas, I., Méndez-Rebolledo, G., Castillo-Retamal, M., Valdés-Badilla, P., & Zapata-Lamana, R. (2020). [Factors associated with low quality of life in Chilean adults during the COVID-19 quarantine]. *Rev Med Chil*, *148*(12), 1759-1766. https://doi.org/10.4067/s0034-98872020001201759
27. Krüger-Malpartida, H., Pedraz-Petrozzi, B., Arevalo-Flores, M., Samalvides-Cuba, F., Anculle-Arauco, V., & Dancuart-Mendoza, M. (2020). Effects on Mental Health After the COVID-19 Lockdown Period: Results From a

Population Survey Study in Lima, Peru. *Clinical Medicine Insights: Psychiatry*, 1-9. https://doi.org/10.1177/1179557320980423

28. López Steinmetz, L. C., Dutto Florio, M. A., Leyes, C. A., Fong, S. B., Rigalli, A., & Godoy, J. C. (2020). Levels and predictors of depression, anxiety, and suicidal risk during COVID-19 pandemic in Argentina: The impacts of quarantine extensions on mental health state. *Psychology, health & medicine*, 1-17. https://doi.org/10.1080/13548506.2020.1867318
29. López Steinmetz, L. C., Godoy, J. C., & Fong, S. B. (2021). A longitudinal study on depression and anxiety in college students during the first 106-days of the lengthy Argentinean quarantine for the COVID-19 pandemic. *Journal of Mental Health*. https://doi.org/10.1080/09638237.2021.1952952
30. López-Morales, H., del Valle, M. V., Canet-Juric, L., Andrés, M. L., Galli, J. I., Poó, F., & Urquijo, S. (2021). Mental health of pregnant women during the COVID-19 pandemic: A longitudinal study. *Psychiatry Research*, *295*. https://doi.org/10.1016/j.psychres.2020.113567
31. Loret de Mola, C., Martins-Silva, T., Carpena, M. X., Del-Ponte, B., Blumenberg, C., Martins, R. C., Dias, I. M., Terribele, F. B., de Avila, A. B., Marmitt, L. P., Meucci, R., & Cesar, J. A. (2021). Maternal mental health before and during the COVID-19 pandemic in the 2019 Rio Grande birth cohort. *Revista brasileira de psiquiatria (Sao Paulo, Brazil : 1999)*. https://doi.org/10.1590/1516-4446-2020-1673
32. Martinez, E. Z., Silva, F. M., Morigi, T. Z., Zucoloto, M. L., Silva, T. L., Joaquim, A. G., DallAgnol, G., Galdino, G., Martinez, M. O. Z., & Silva, W. R. da. (2020). Physical activity in periods of social distancing due to COVID-19: A cross-sectional survey^ienAtividade física em períodos de distanciamento social

devidos à COVID-19: Um estudo transversal^ipt. *Ciênc. Saúde Colet*, *25*(supl.2), 4157-4168.

33. Meda-Lara, R. M., Muñoz-Valle, J. F., Juárez-Rodríguez, P., Figueroa-López, C., Herrero, M., de Santos Ávila, F., Palomera-Chávez, A., Yeo Ayala, C., & Moreno-Jiménez, B. (2021). Psychological responses to COVID-19 in a Mexican population: An exploratory study during second and third phases. *Psychology, health & medicine*, 1-8. https://doi.org/10.1080/13548506.2021.1883689
34. Miranda, A. R., Scotta, A. V., Cortez, M. V., & Soria, E. A. (2021). Triggering of postpartum depression and insomnia with cognitive impairment in Argentinian women during the pandemic COVID-19 social isolation in relation to reproductive and health factors. *Midwifery*, *102*, 103072. https://doi.org/10.1016/j.midw.2021.103072
35. Montero Doig, A. M., Torres, G. Y., & Correa-Lopez, L. E. (2021). Resilience, anxiety and eating habits of the south-east amazon population before and during the pandemic. *Revista de la Facultad de Medicina Humana*, *21*(3), 557-563. https://doi.org/10.25176/RFMH.V21I3.3732
36. Monterrosa-Blanco, A., Monterrosa-Castro, Á., & González-Sequeda, A. (2021). Online assessment of the perception of loneliness and associated factors in Colombian climacteric women during the COVID-19 pandemic: A cross-sectional study. *Health Promotion Perspectives*, *11*(2), 230-239. https://doi.org/10.34172/hpp.2021.28
37. Monterrosa-Castro, Á., Monterrosa-Blanco, A., & González-Sequeda, A. (2021). Perceived Loneliness and Severe Sleep Disorders in Adult Women during the Covid-19 Quarantine: A Cross-Sectional Study in Colombia. *Journal*

*of Primary Care & Community Health*, 1-8. https://doi.org/10.1177/21501327211025170

38. Naranjo-Hernández, Y., Mayor-Walton, S., Rivera-García, O. de la, & González-Bernal, R. (2021). Estados emocionales de adultos mayores en aislamiento social durante la COVID-19^iesEmotional states of older adults in social isolation during COVID-19^ienEstados emocionais de idosos em isolamento social durante COVID-19^ipt. *Rev. inf. cient*, *100*(2). es
39. Nomura, R., Tavares, I., Ubinha, A. C., Costa, M. L., Opperman, M. L., Brock, M., Trapani, A., Damasio, L., Reis, N., Borges, V., Zaconeta, A., Araujo, A. C., & Ruano, R. (2021). Impact of the covid-19 pandemic on maternal anxiety in Brazil. *Journal of Clinical Medicine*, *10*(4), 1-14. https://doi.org/10.3390/jcm10040620
40. Oblitas González Correo, A., & Sempertegui Sánchez Correo, N. (2020). Ansiedad en tiempos de aislamiento social por COVID-19. Chota, Perú, 2020. *Avances en Enfermeria*, *38*, 11-21. https://doi.org/10.15446/av.enferm.v38n2.87589
41. Paico, N. I. H., Sebastian, S. D., Vergaray, J. M., Arellano, E. G. R., & Navarro, E. R. (2021). Anxiety and aggressiveness in Peruvian postgraduate students in COVID-19 context. *European Journal of Molecular and Clinical Medicine*, *8*(3), 713-726.
42. Passos, L., Prazeres, F., Teixeira, A., & Martins, C. (2020). Impact on mental health due to covid-19 pandemic: Cross-sectional study in portugal and brazil. *International Journal of Environmental Research and Public Health*, *17*(18), 1-13. https://doi.org/10.3390/ijerph17186794

52. Riter, H. da S., Almeida, M. L., Vescovi, G., Marques, F. M., Pedrotti, B. G., Mallmann, M. Y., Pieta, M. A. M., & Frizzo, G. B. (2021). Symptoms of common mental disorders in brazilian parents during the covid-19 pandemic: Associated factors. *Psychological Studies*, No-Specified. https://doi.org/10.1007/s12646-021-00609-8
53. Ruiz-Frutos, C., Palomino-Baldeón, J. C., Ortega-Moreno, M., Villavicencio-Guardia, M. D. C., Dias, A., Bernardes, J. M., & Gómez-Salgado, J. (2021). Effects of the COVID-19 Pandemic on Mental Health in Peru: Psychological Distress. *Healthcare (Basel)*, *9*(6). https://doi.org/10.3390/healthcare9060691
54. Schmitt, A. A., Brenner, A. M., Primo de Carvalho Alves, L., Claudino, F. C. de A., Fleck, M. P. de A., Rocha, N. S., & Schmitt, A. A., Jr. (2021). Potential predictors of depressive symptoms during the initial stage of the COVID-19 outbreak among Brazilian adults. *Journal of Affective Disorders*, *282*, 1090-1095. https://doi.org/10.1016/j.jad.2020.12.203
55. Scotta, A. V., Cortez, M. V., & Miranda, A. R. (2020). Insomnia is associated with worry, cognitive avoidance and low academic engagement in Argentinian university students during the COVID-19 social isolation. *Psychology, health & medicine*, 1-16. https://doi.org/10.1080/13548506.2020.1869796
56. Seco Ferreira, D. C., Oliveira, W. L., Costa Delabrida, Z. N., Faro, A., & Cerqueira-Santos, E. (2020). Intolerance of uncertainty and mental health in Brazil during the Covid-19 pandemic^ienIntolerância a incerteza e saúde mental no Brasil durante a pandemia de Covid-19^ipt. *Suma psicol*, *27*(1), 62-69.
57. Serafim, A. P., Durães, R. S. S., Rocca, C. C. A., Gonçalves, P. D., Saffi, F., Cappellozza, A., Paulino, M., Dumas-Diniz, R., Brissos, S., Brites, R., Alho, L., & Lotufo-Neto, F. (2021). Exploratory study on the psychological impact of

COVID-19 on the general Brazilian population. *PLoS ONE*, *16*(2 February). https://doi.org/10.1371/journal.pone.0245868

58. Silva, L. R. B., Seguro, C. S., de Oliveira, C. G. A., Santos, P. O. S., de Oliveira, J. C. M., de Souza Filho, L. F. M., de Paula Júnior, C. A., Gentil, P., & Rebelo, A. C. S. (2020). Physical Inactivity Is Associated With Increased Levels of Anxiety, Depression, and Stress in Brazilians During the COVID-19 Pandemic: A Cross-Sectional Study. *Frontiers in Psychiatry*, *11*. https://doi.org/10.3389/fpsyt.2020.565291
59. Soares, L. L., Leite, L. B., Guilherme, L. Q., Rezende, L. M., Noce, F., & Pussieldi, G. (2021). Anxiety, sleep quality and mood in elite athletes during the COVID-19 pandemic: A preliminary study. *J Sports Med Phys Fitness*. https://doi.org/10.23736/s0022-4707.21.12276-5
60. Souza, A. S. R., Souza, G. F. A., Souza, G. A., Cordeiro, A. L. N., Praciano, G. A. F., Alves, A. C. S., Santos, A. C. D., Silva Junior, J. R., & Souza, M. B. R. (2021). Factors associated with stress, anxiety, and depression during social distancing in Brazil. *Revista de saude publica*, *55*, 5. https://doi.org/10.11606/s1518-8787.2021055003152
61. Stults-Kolehmainen, M., Filgueiras, A., & Blacutt, M. (2021). Factors linked to changes in mental health outcomes among Brazilians in quarantine due to COVID-19. *medRxiv*, 2020.05.12.20099374. https://doi.org/10.1101/2020.05.12.20099374
62. Suárez-Rico, B. V., Estrada-Gutierrez, G., Sánchez-Martínez, M., Perichart-Perera, O., Rodríguez-Hernández, C., González-Leyva, C., Osorio-Valencia, E., Cardona-Pérez, A., Helguera-Repetto, A. C., Sosa, S. E. Y., Solis-Paredes, M., & Reyes-Muñoz, E. (2021). Prevalence of depression, anxiety, and perceived

stress in postpartum mexican women during the covid-19 lockdown. *International Journal of Environmental Research and Public Health*, *18*(9). https://doi.org/10.3390/ijerph18094627

63. Terán-Pérez, G., Portillo-Vásquez, A., Arana-Lechuga, Y., Sánchez-Escandón, O., Mercadillo-Caballero, R., González-Robles, R. O., & Velázquez-Moctezuma, J. (2021). Sleep and mental health disturbances due to social isolation during the covid-19 pandemic in Mexico. *International Journal of Environmental Research and Public Health*, *18*(6), 1-11. https://doi.org/10.3390/ijerph18062804
64. Toledo-Fernández, A., Betancourt-Ocampo, D., & González-González, A. (2021). Distress, Depression, Anxiety, and Concerns and Behaviors Related to COVID-19 during the First Two Months of the Pandemic: A Longitudinal Study in Adult MEXICANS. *Behav Sci (Basel)*, *11*(5). https://doi.org/10.3390/bs11050076
65. Torales, J., Ríos-González, C., Barrios, I., O'Higgins, M., González, I., García, O., Castaldelli-Maia, J. M., & Ventriglio, A. (2020). Self-Perceived Stress During the Quarantine of COVID-19 Pandemic in Paraguay: An Exploratory Survey. *Frontiers in Psychiatry*, *11*. https://doi.org/10.3389/fpsyt.2020.558691
66. Torrente, F., Yoris, A., Low, D. M., Lopez, P., Bekinschtein, P., Manes, F., & Cetkovich, M. (2021). Sooner than you think: A very early affective reaction to the COVID-19 pandemic and quarantine in Argentina. *Journal of Affective Disorders*, *282*, 495-503. https://doi.org/10.1016/j.jad.2020.12.124
67. Torrente, F., Yoris, A., Low, D. M., Lopez, P. L., Bekinschtein, P., Vázquez, G., Manes, F., & Cetkovich, M. (2021). Emotional symptoms, mental fatigue and behavioral adherence after 72 continuous days of strict lockdown during the

COVID-19 pandemic in Argentina. *medRxiv*, 2021.04.21.21255866. https://doi.org/10.1101/2021.04.21.21255866

68. Tyler, C. M., McKee, G. B., Alzueta, E., Perrin, P. B., Kingsley, K., Baker, F. C., & Arango-Lasprilla, J. C. (2021). A study of older adults' mental health across 33 countries during the covid-19 pandemic. *International Journal of Environmental Research and Public Health*, *18*(10). https://doi.org/10.3390/ijerph18105090
69. Vitorino, L. M., Yoshinari Júnior, G. H., Gonzaga, G., Dias, I. F., Pereira, J. P. L., Ribeiro, I. M. G., França, A. B., Al-Zaben, F., Koenig, H. G., & Trzesniak, C. (2021). Factors associated with mental health and quality of life during the COVID-19 pandemic in Brazil. *BJPsych Open*, *7*(3). https://doi.org/10.1192/bjo.2021.62
70. Vizioli, N. A., & Crespi, M. (2021). Factores estresantes y sintomatología psicológica durante el Aislamiento Social Preventivo Obligatorio por COVID-19 en población adulta de Buenos Aires / Stressors and Psychological Symptoms during Social, Preventive and Mandatory Isolation for COVID-19. *Subj. procesos cogn*, *24*(2), 17-41.
71. Zhang, S. X., Wang, Y., Jahanshahi, A. A., Li, J., & Schmitt, V. G. H. (2021). Early evidence and predictors of mental distress of adults one month in the COVID-19 epidemic in Brazil. *Journal of Psychosomatic Research*, *142*, N.PAG-N.PAG. https://doi.org/10.1016/j.jpsychores.2021.110366
